# Multimodal classification of molecular subtypes in pediatric acute lymphoblastic leukemia

**DOI:** 10.1101/2023.03.24.23287613

**Authors:** Olga Krali, Yanara Marincevic-Zuniga, Gustav Arvidsson, Anna Pia Enblad, Anders Lundmark, Shumaila Sayyab, Vasilios Zachariadis, Merja Heinäniemi, Janne Suhonen, Laura Oksa, Kaisa Vepsäläinen, Ingegerd Öfverholm, Gisela Barbany, Ann Nordgren, Henrik Lilljebjörn, Thoas Fioretos, Hans O. Madsen, Hanne Vibeke Marquart, Trond Flaegstad, Erik Forestier, Ólafur G Jónsson, Jukka Kanerva, Olli Lohi, Ulrika Norén-Nyström, Kjeld Schmiegelow, Arja Harila, Mats Heyman, Gudmar Lönnerholm, Ann-Christine Syvänen, Jessica Nordlund

**Author notes:** Corresponding author: Dr. Jessica Nordlund, Box 1432, BMC, 75144 Uppsala, Sweden.

## Abstract

Genomic analyses have redefined the molecular subgrouping of pediatric acute lymphoblastic leukemia (ALL). Molecular subgroups guide risk-stratification and targeted therapies, but outcomes of recently identified subtypes are often unclear, owing to limited cases with comprehensive profiling and cross-protocol studies. We developed a machine learning tool (ALLIUM) for the molecular subclassification of ALL in retrospective cohorts as well as for up-front diagnostics. ALLIUM uses DNA methylation and gene expression data from 1131 Nordic ALL patients to predict 17 ALL subtypes with high accuracy. ALLIUM was used to revise and verify the molecular subtype of 280 cases with undefined/B-other molecular phenotype, resulting in a single revised subtype for 85.4% of these cases. Our study shows the power of combining DNA methylation and gene expression data for resolving ALL subtypes and provides the first comprehensive population-based retrospective cohort study of molecular subtype frequencies in the Nordic countries, identifying subgroups with differential survival outcomes.

## Introduction

Pediatric acute lymphoblastic leukemia (ALL) comprises a heterogeneous group of patients who can be stratified into subgroups based on the presence of recurrent cytogenetic aberrations, which are important predictors of clinical outcome^1,2^. Subtypes of pediatric B-cell precursor ALL (BCP-ALL) are often characterized by large-scale chromosomal aberrations, including abnormal chromosomal numbers, translocations that give rise to expressed fusion genes, or other structural rearrangements. Before next-generation sequencing (NGS)-based methods were introduced into clinical practice, as many as 30% of all BCP-ALL cases lacked conclusive results from standard cytogenetic analyses (denoted B-other) and therefore subtype information was not available for treatment stratification or disease monitoring^3^. Recent application of high-resolution transcriptome sequencing (RNA-seq) has enabled the discovery of new oncogenic subgroups characterized by fusion genes, such as *DUX4* (*DUX4*-r), *ZNF384* (*ZNF384*-r), *MEF2D* (*MEF2D*-r) and *NUTM1* (*NUTM1*-r) rearrangements^4–15^, as well as subtype-like signatures, such as *BCR::ABL1*-like/”Ph-like”^16,17^ or *ETV6::RUNX1*-like/”ER-like”^7,18^, and the PAX5-driven subtypes, PAX5 alteration (PAX5alt) and PAX5 P80R^19–22^. The clinical significance of the recently identified subtypes is often unclear, owing to the limited number of cases and differences between protocols and studies^7,8,20,23,24^. Therefore, retrospective ALL cohort analyses have been particularly powerful for studying rare ALL subtypes due to the large sample sizes available in biobanks and the prolonged period of follow-up to collect sufficient data on rare events

Most of the recurrent molecular alterations in ALL are strongly associated with gene expression (GEX) profiles^24,25^. RNA-seq has since emerged as a powerful tool for the identification of both fusion genes and GEX subtype profiling in a single assay^26,27^, which promises to replace cumbersome standard karyotyping (G-banding), PCR-based and fluorescence in situ hybridization (FISH)-based methods in a clinical diagnostic setting^28^. Compared to DNA, RNA is prone to degradation, making it challenging to obtain high-quality RNA for retrospective cohort analyses. However, epigenetic profiling of DNA methylation (DNAm) using arrays or next-generation sequencing (NGS) has demonstrated comparable subtype-specific distributions in ALL cells^29–32^. DNAm is advantageous as an analyte due to its ability to identify methylation patterns associated with disease in degraded archival samples^33^. Leveraging biobank samples and retrospective cohort studies can provide valuable insights into long-term disease outcomes that may be challenging to obtain through prospective study designs, particularly for rare ALL subtypes.

In the present investigation, we describe a multimodal machine learning classification tool, <u>ALL</u> subtype Identification <u>U</u>sing <u>M</u>achine learning (ALLIUM) that uses DNAm and/or GEX signatures (**Figure 1**). We trained and applied ALLIUM to a large cohort of 1131 Nordic patient samples and determined the frequencies of recent genetic subtypes, which led to the revision of molecular subtypes in 85.4% of B-other cases.

**Figure 1.**
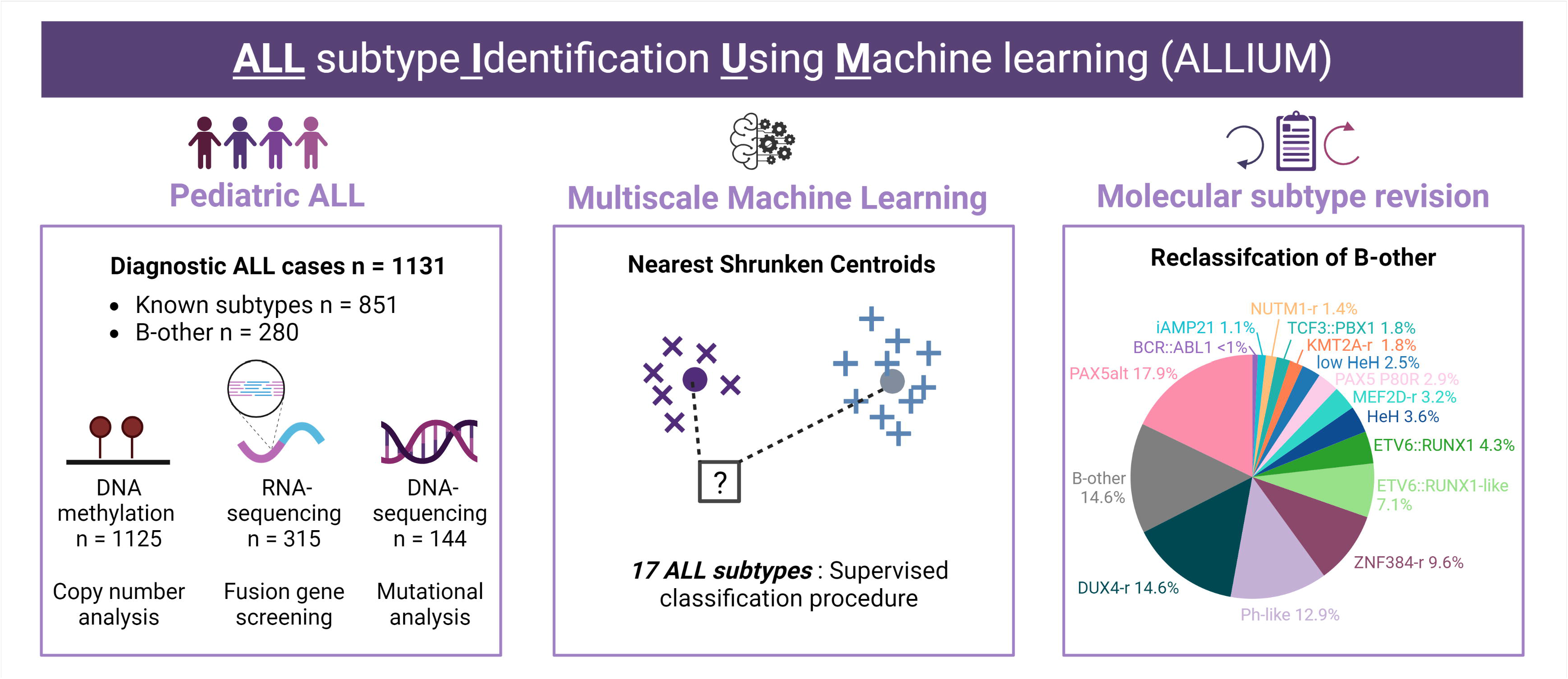
Study overview. DNA methylation (DNAm, 450k arrays), gene expression (GEX, RNA-sequencing), and somatic mutation (WGS, targeted sequencing) data were generated from 1131 patients treated on the Nordic Society for Pediatric Hematology and Oncology (NOPHO) protocols diagnosed between 1996 and 2013. Molecular subtyping was performed based on a combination of cytogenetics, fusion gene screening, mutational analysis, and copy number analysis. A supervised classification method was used to build subtype-specific models based on DNAm and GEX profiles for 17 molecular subtypes of ALL. B-other patients were reclassified using multimodal subtype classification with ALLIUM.

## Results

### Molecular characteristics and data generation

Diagnostic bone marrow aspirates or peripheral blood samples from 1131 Nordic ALL patients (n = 1025 BCP-ALL and n = 106 T-ALL) were obtained from a population based cohort diagnosed between 1996 and 2013, and enrolled in the Nordic Society of Pediatric Hematology and Oncology (NOPHO) -92, - 2000, -2008, EsPh-ALL, or Interfant treatment protocols^34–37^. Genome-wide CpG methylation levels were analyzed in 1125 DNA samples (1125 patients) using 450k arrays (DNAm dataset) and RNA-sequencing was performed in 328 RNA samples (315 patients, GEX dataset). Molecular subtypes were assigned based on standard cytogenetic analysis at ALL diagnosis, where a total of 851 patients (75%) had an established molecular subtype and 280 were denoted B-other (Supplementary Table S1). We initially screened our cohort for the molecular ALL subtypes outlined by the International Consensus Classification (ICC)^2^ using a combination of genome-wide CNA detection, fusion gene screening (Supplementary Figure S1), and targeted mutational assessment for *PAX5* p.Pro80Arg, *IKZF1* p.Asn159Tyr, and *ZEB2* p.His1038Arg (Supplementary Table S2). This analysis, combined with putative revised molecular subtype information from previously published results^7,9,30,38–40^, identified 131 unique fusion genes in 225 patients, and 130 patients from the B-other group (46%) who belonged to one of the ICC subtypes. In total, this yielded 981 ICC subtype-defined cases, while 150 cases remained as B-other (**Table 1**). Of note, this included 27 patients with established subtypes missed by routine diagnostics: HeH (n = 9), *ETV6::RUNX1* (n = 9), *KMT2A*-r (n = 4), *TCF3::PBX1* (n = 3), *BCR::ABL1* (n = 1), and iAMP21 (n = 1). One patient (ALL_913) was re-labelled from HeH to *DUX4*-r, after confirmation of the presence of the *IGH-DUX4* fusion gene and a normal copy number. The 30 patients with dic(9;20) aberrations^41^ were classified as PAX5alt.

**Table 1.** Overview of the 1131 ALL patients by ICC subtype prior to multimodal classification.

| Molecular subtype* | # Patients<br>DNAm | # Patients<br>GEX | # Patients<br>Total | Age (sd) | # Male/Female |
| --- | --- | --- | --- | --- | --- |
| Total patients | 1125 | 315 | 1131 | 6.35 (± 4.39) | 615/516 |
| ICC subtype-defined | 975 | 251 | 981 | 6.03 (± 4.2) | 534/447 |
| B-other | 150 | 64 | 150 | 8.45 (± 4.99) | 81/69 |
| **HeH** | 309 | 46 | 310 | 5.11 (± 3.51) | 166/144 |
| **low HeH** | 5 | 3 | 5 | 3.27 (± 1.33) | 2/3 |
| **iAMP21** | 20 | 16 | 21 | 10.23 (± 3.95) | 13/8 |
| **Hypodiploidy** | 11 | 0 | 11 | 8.97 (± 4.71) | 6/5 |
| ***ETV6::RUNX1*** | 274 | 32 | 275 | 4.93 (± 2.55) | 144/131 |
| ***ETV6::RUNX1*-like** | 12 | 10 | 12 | 4.13 (± 3.66) | 5/7 |
| **T-ALL** | 105 | 19 | 106 | 9.08 (± 4.61) | 78/28 |
| ***KMT2A-r*** | 61 | 14 | 62 | 2.76 (± 4.22) | 26/36 |
| ***NUTM1-r*** | 3 | 3 | 3 | 9.46 (± 7.78) | 1/2 |
| **PAX5alt** | 52 | 33 | 53 | 5.93 (± 5.07) | 25/28 |
| **PAX5 P80R** | 5 | 4 | 5 | 12.35 (± 5.83) | 4/1 |
| ***TCF3::PBX1*** | 37 | 10 | 37 | 8.07 (± 4.55) | 17/20 |
| ***MEF2D-r*** | 9 | 8 | 9 | 11.3 (± 4.57) | 3/6 |
| ***BCR::ABL1*** | 25 | 10 | 25 | 8.92 (± 3.82) | 15/10 |
| ***BCR::ABL1*-like** | 10 | 7 | 10 | 8.82 (± 5.75) | 8/2 |
| ***DUX4-r*** | 20 | 19 | 20 | 9.72 (± 3.11) | 11/9 |
| ***ZNF384-r*** | 17 | 17 | 17 | 9.02 (± 3.78) | 10/7 |

\* Molecular subtypes were labelled according to the International Consensus Classification (ICC).
Fluorescence in situ hybridization and/or reverse-transcriptase polymerase chain reaction were applied at ALL diagnosis to identify established subtypes *ETV6::RUNX1*, *TCF3::PBX1*, *KMT2A-r*, dic(9;20), iAMP21. High hyperdiploidy (HeH) was defined as a modal number ≥ 51 chromosomes or DNA Index (DI) 1.12-1.35. Hypodiploidy was defined as $<$ 40 chromosomes and included low-hypodiploidy with 30-39 chromosomes or DI 0.6-0.84 and near-haploidy (NH) with 24-29 chromosomes or DI $<$ 0.6. Low HeH was defined as 48-50 chromosomes as determined by array-based copy number analysis. All karyotypes were centrally reviewed.

### ALLIUM is a highly sensitive method for molecular ALL subtype classification

In order to design a DNAm and GEX-based classifiers for ALL, the 981 patients with known ICC molecular subtypes defined based on updated molecular analysis were split into design and hold-out datasets to create and validate the ALLIUM classifier (**Table 2**). An internally produced replication set (n = 13, GEX) and three external datasets (GEX: GSE161501^42^, GEX NOPHO-Finland and DNAm: GSE56600^43^) were used for additional independent subtype verification. ALLIUM is based on nearest shrunken centroid (NSC) models consisting of DNAm and GEX data in a one vs. rest approach^44^. Subtypes with similar molecular profiles, i.e. those characterized by aneuploidies (HeH, low HeH, iAMP21, hypodiploidy), *ETV6* gene rearrangements (*ETV6::RUNX1, ETV6::RUNX1*-like), and the Philadelphia (ph) chromosome (*BCR::ABL1, BCR::ABL1*-like) were handled in a different manner. For these subtypes, a two-step procedure with initial classification on the group level, followed by a one-vs-one or a multi-class classification within the group was applied (Supplementary Materials and Methods and Supplementary Figure S2–3). Moreover, to identify misclassification errors due to low blast count, control classifiers for DNAm and RNA were built utilizing data available from ALL patients in remission or healthy blood donors^45^. As the output contained probability scores for each classifier, multiple subtype classifications could occur. Therefore, we proceeded with a multi- to single-class transformation, assigning the subtype with the highest probability score for each sample.

**Table 2.**
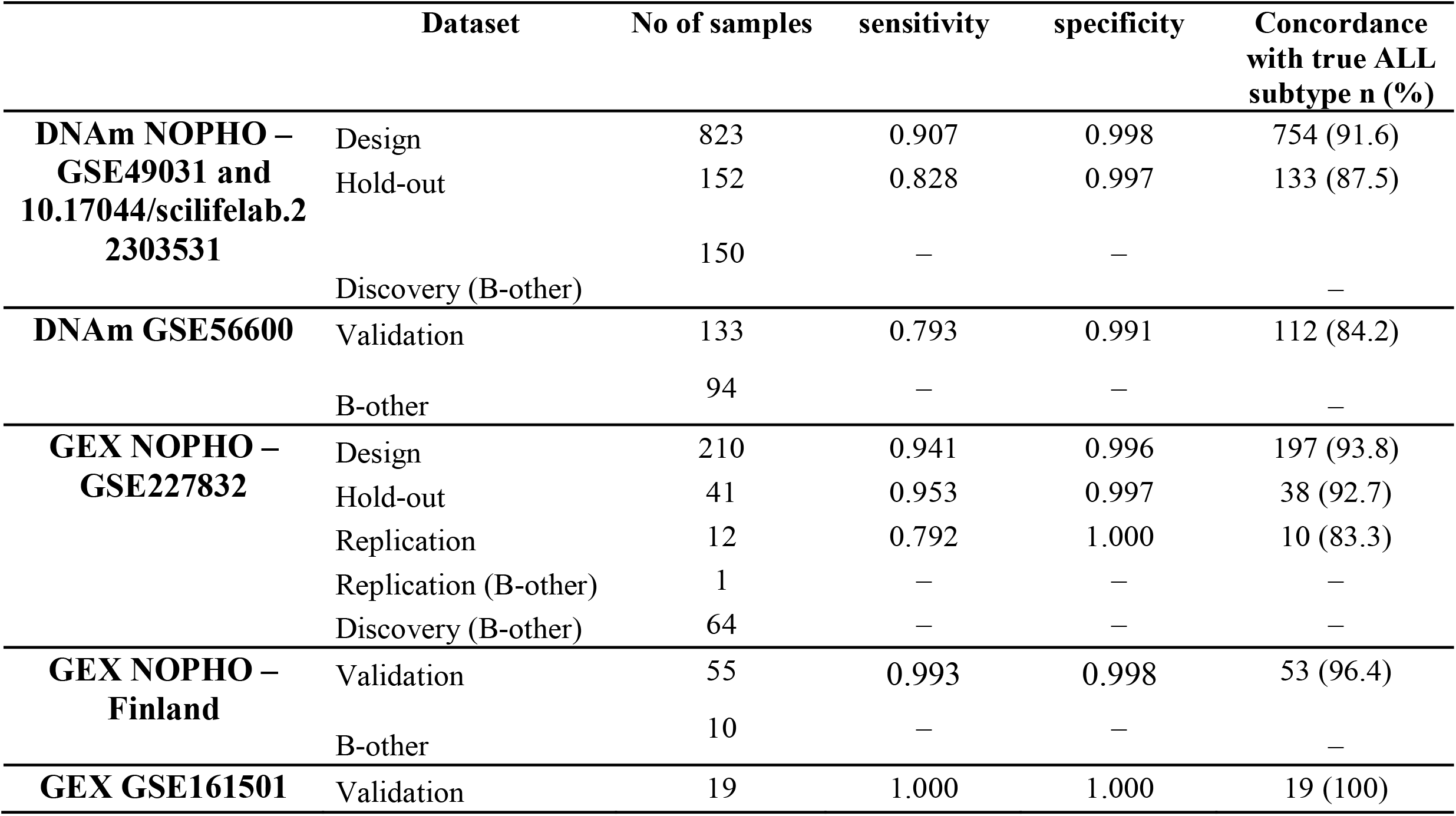
Classifier performance and concordance.

ALLIUM identified 519 CpGs and 425 genes as most informative for subtype determination (Supplementary Table S3–4). Unsupervised analysis of samples with known subtype revealed clear subtype-driven clustering (**Figure 2a-c**, Supplementary Figure S4). We evaluated the models using hold-out, replication and independent external validation datasets (**Table 2**, Supplementary Table S5–13). The classifiers were highly predictive overall, with 86.0% concordance between DNAm and true molecular subtype and 94.5% overall concordance between GEX and true molecular subtype (**Figure 2b-d, Table 2**). Both DNAm and GEX classification was performed in 245 patients in our dataset, and the classifiers achieved 92.2% overall concordance (design: 92.6%, 189/204, hold-out: 90.2%, 37/41, **Figure 2e-f**). However, the performance varied between the subtypes (**Figure 2g**, Supplementary Table S14). In total, only 19 cases showed discrepant result between the GEX and DNAm classifier, where the GEX alone was correct for eight, the DNAm was correct for six, and complete mismatches were observed for the remaining five cases (Supplementary Table S15). Of the eight patients that were correctly predicted by GEX, but not by DNAm, five were predicted as no-class by the DNAm classifier. Overall, ALLIUM DNAm was correct for 93.5%, ALLIUM GEX was correct for 94.3%, and with both modalities available a correct classification as achieved in 96.7% (Supplementary Table S16). Additional details about the performance of the replication and external validation datasets can be found in the **supplementary results** (Supplementary Figure S4).

**Figure 2.**
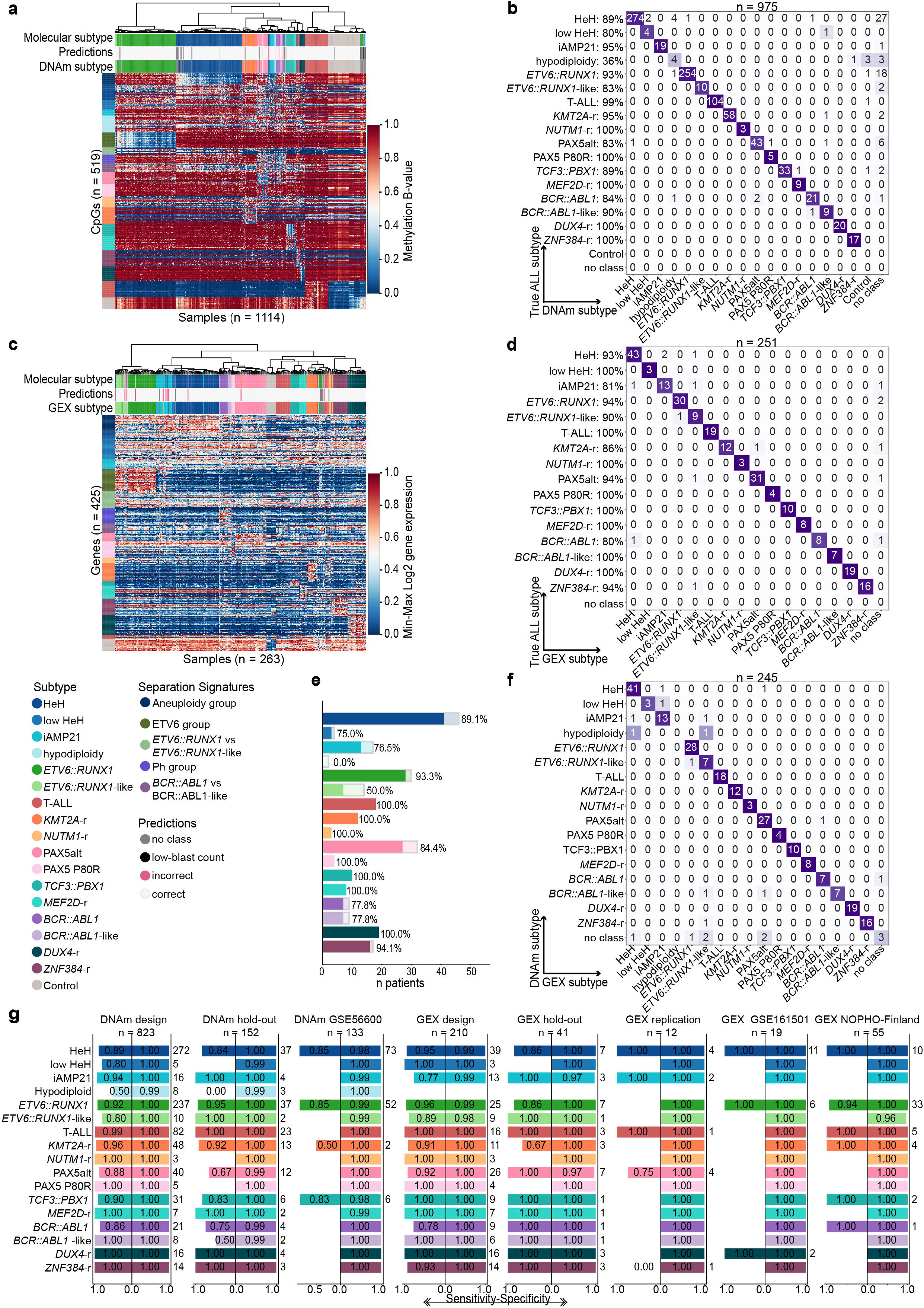
Evaluation of model performance. a) Unsupervised hierarchical clustering based on DNA methylation (DNAm) levels of 519 CpG sites across molecularly defined patients (n = 975) and control samples (n = 139). b) Concordance between ALLIUM DNAm subtype predictions (x-axis) and true molecular subtypes (y-axis) for the 975 patients. c) Unsupervised hierarchical clustering based on gene expression (GEX) levels of 425 genes across molecularly defined patients (n = 251) and control samples (n = 12). d) Concordance between ALLIUM GEX subtype predictions (x-axis) and true molecular subtypes (y-axis) for the 251 patients. e) The degree of concordance between DNAm and GEX predictions (“no class” predictions are not shown). The light bars represent the overall predictions per subtype and the darker bars indicate the number of concordant predictions. f) Concordance of DNAm (x-axis) and GEX (y-axis) subtype predictions (n = 245 patients). g) Sensitivity and specificity across 17 ALL subtypes for our design, hold-out and replication datasets, and the three external validation datasets, DNAm GSE56600, GEX GSE161501 and GEX NOPHO-Finland.

### Functional annotation of genes and CpG sites identified by ALLIUM

Next we annotated the 519 CpG sites and 425 genes selected by ALLIUM. In general, the subtype-defining signatures were more often characterized by hypomethylation (Supplementary Figure S5) and increased, but also variable gene expression (Supplementary Figure S6). We studied the intersection of the genomic location of the CpG sites and genes selected by ALLIUM. A total of 31 CpG sites overlapped with the genomic location of 19 of the genes selected by the ALLIUM GEX (Supplementary Table S17-18). Among these, 22 CpG sites located in 12 genes were selected for the same subtype i.e. *ETV6::RUNX1* vs *ETV6::RUNX1*-like with 5 CpG sites in *MGC70857/C8orf82* and *ARHGEF12*, the *ETV6* group with 4 CpG sites in *IGF2BP1* and *FARP1*, the aneuploidy group with 4 CpG sites in *S100A16* and *FUT7*, and the Philadelphia group with 3 CpG sites in *CA6*, and differentiating *BCR::ABL1* from *BCR:ABL1*-like with 2 CpG sites in *SPRED2*. For these genes we observed an inverse relationship between methylation level and gene expression (Pearson’s correlation coefficient = -0.64, Supplementary Figure S7).

The genomic features selected by ALLIUM were distributed across all chromosomes, without a significant overall enrichment in the location of CpG sites or genes (FDR q-value > 0.05, Supplementary Figure S8 and Supplementary Table S19-20), although the CpGs sites were significantly enriched outside CpG islands in intergenic “open sea” regions (FDR q-value < 0.0001, Supplementary Table S21). The distribution of CpG sites varied among the subtypes. For example, a significant overrepresentation (44%, FDR q-value = 0.004) of the CpG sites differentiating *ETV6::RUNX1* from *ETV6::RUNX1*-like, were annotated on chromosome 8 in the vicinity of the *MGC70857*/*C8orf82* and *ANK1* loci and *MGC70857* was selected by the GEX classifier. About 25% of the genes selected by the *KMT2A*-r GEX classifier were located on chromosome 7 in the HOX gene cluster (FDR q-value 0.02). Additionally, the GEX classifier selected *NUTM1* for *NUTM1*-r, *PBX1* for *TCF3::PBX1, ABL1* for differentiating *BCR::ABL1* from *BCR::ABL1*-like, and *MEF2C* for *MEF2D*-r (**Figure 3a**). The classifiers selected additional biologically relevant genes and CpG sites for ALL, including CpG sites in *CBFA2T3* and *EPOR* for the *ETV6*-group, expression of *CDKN2A* and CpG sites in *AUTS2* for the PAX5alt group, expression of *CEBPA* for *ZNF384*-r, and CpG sites in *ETV6, RUNX2*, and *IKZF1* for PAX5 P80R (**Figure 3b**).

**Figure 3.**
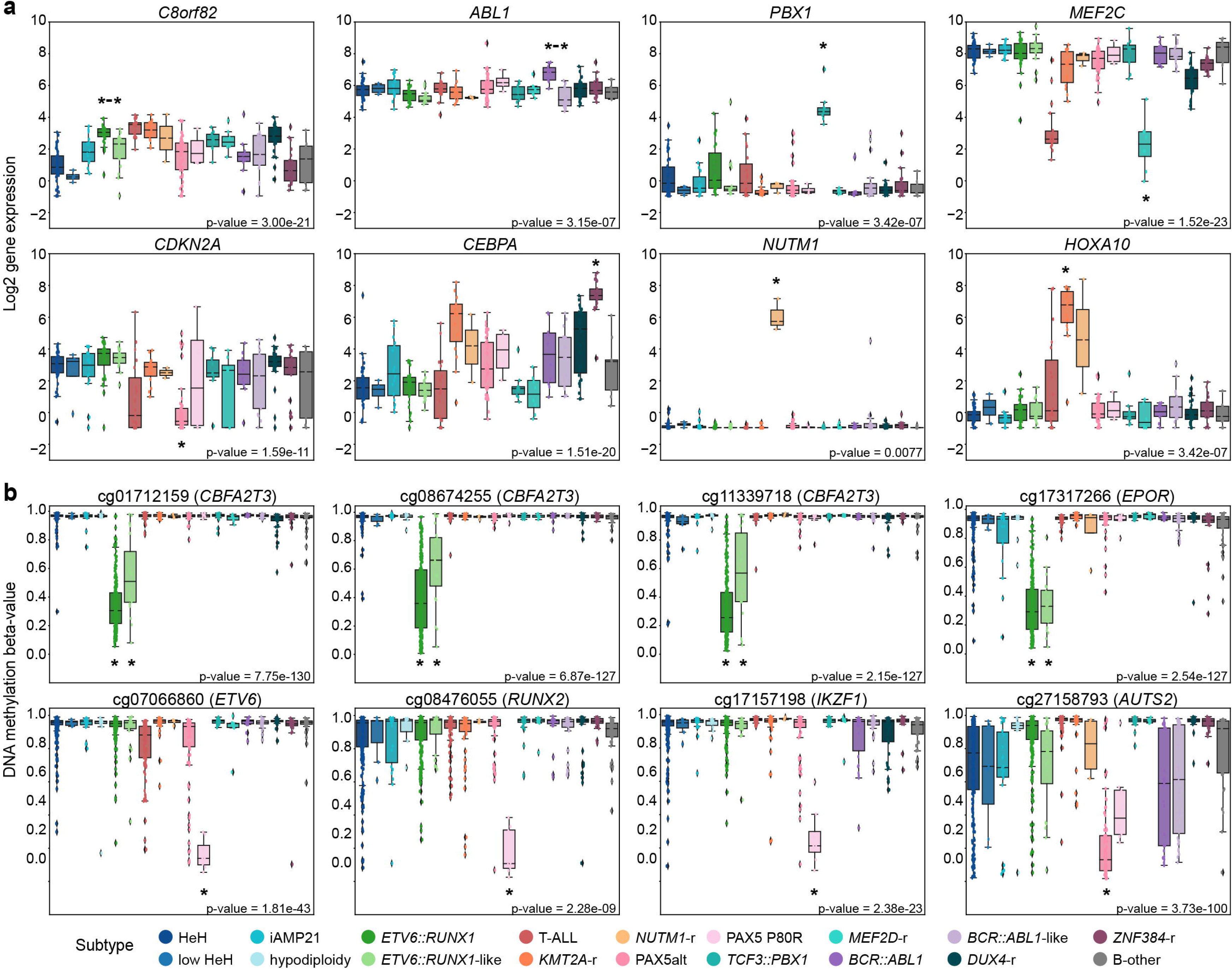
Subtype-specific signatures determined by ALLIUM. Boxplots demonstrating the a) GEX levels for eight genes across 315 patients grouped by revised molecular subtype. b) DNAm levels for eight CpG sites across 1125 patients by revised molecular subtype. The boxes are color-coded by respective subtype according to the key at the bottom. The Benjamini-Hochberg (BH) corrected Kruskal-Wallis H-test p-value indicates the statistical significance between subtypes (bottom right). Asterisks indicate the subtype(s) for which ALLIUM chose each specific CpG or GEX signature, while connected asterisks indicate signatures selected to differentiate subtypes within larger groups.

### Comparisons of GEX model performance

We further evaluated the ALLIUM GEX classifier against two other GEX classifiers for BCP-ALL; ALLSorts and ALLCatchR^26,27^. As both classifiers are trained specifically for BCP-ALL, we removed T-ALL from the comparison. ALLIUM GEX, ALLSorts, and ALLCatchR were evaluated for the 312 BCP-ALL samples of known subtype across all the five GEX datasets included herein. Overall, the three classifiers performed similarly (**Figure 4a-c**), although notable differences between ALLIUM GEX, ALLSorts, and ALLCatchR, respectively, were observed for classification of PAX5alt (100%, 71%, 71%), HeH (86%, 71%, 86%), and iAMP21 (100%, 33%, 67%) using our hold-out dataset (Supplementary Figures S9-12). Specifically, we noted that ALLSorts and ALLCatchR predicted 3 (including a multi-class case) and 8 PAX5alt patients with dic(9;20) (n = 14) as ph-like, respectively. ALLIUM was not trained on *BCL2/MYC, IKZF1 N159Y, HLF, CEB* and *CDX2/UBTF* subtypes and ALLSorts and ALLCatchR did not predict any of these rare subtypes in our cohort.

**Figure 4.**
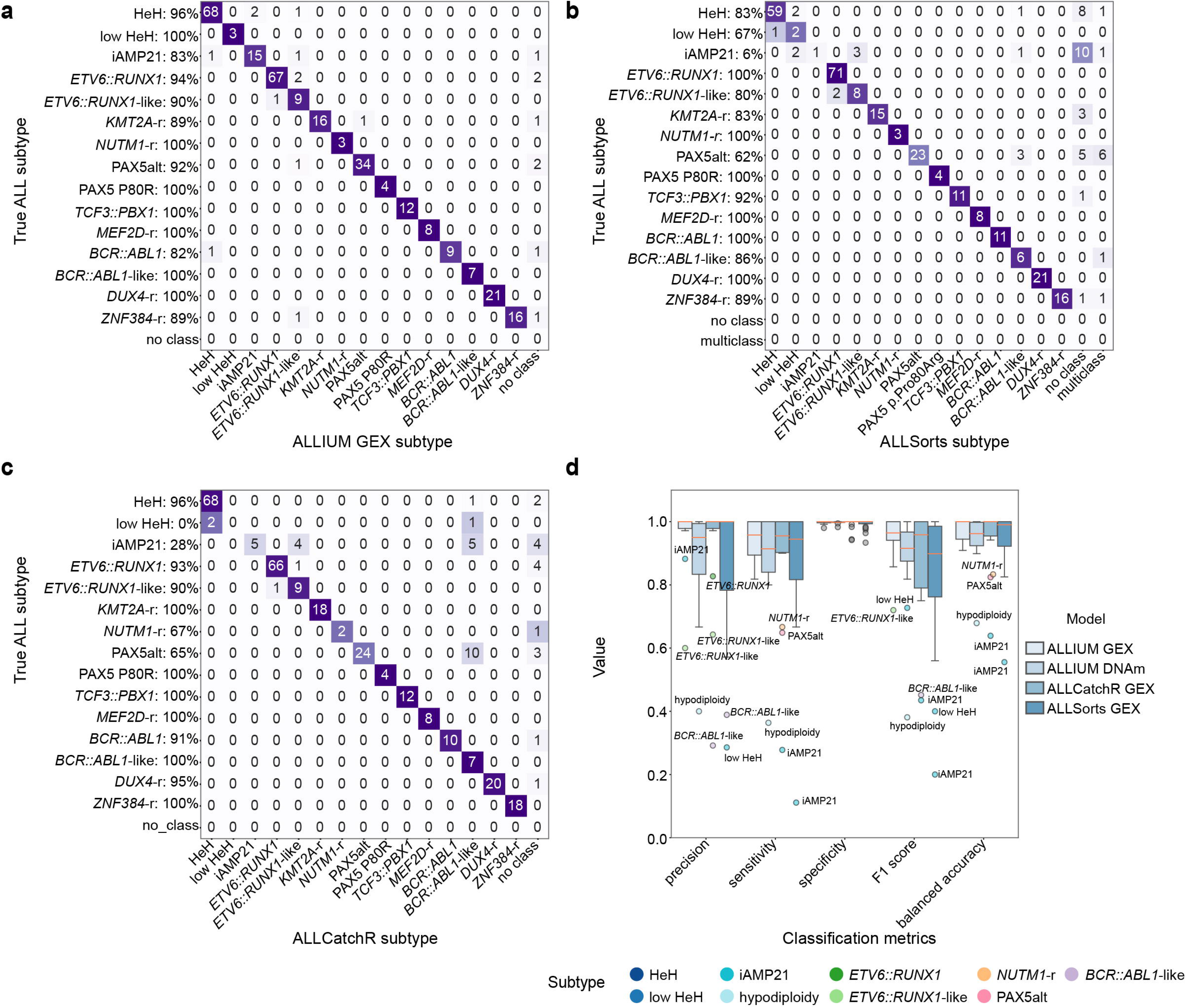
Performance of ALLIUM, ALLSorts and ALLCatchR. a) Concordance between ALLIUM GEX subtype predictions (x-axis) and true molecular subtypes (y-axis) for 312 BCP-ALL samples of known subtype. b) Concordance between ALLSorts subtype predictions (x-axis) and true molecular subtype (y-axis). c) Concordance between ALLCatchR subtype predictions (x-axis) and true molecular subtype (y-axis). d) Boxplots demonstrating classification performance, including precision, sensitivity, specificity, F1 score and accuracy (balanced) for the three GEX models (n = 312 samples) and ALLIUM DNAm (n = 1108 samples with known subtype).

ALLIUM DNAm performed similarly to the GEX models (**Figure 4d**). No other model is currently available for subtyping in ALL by DNA methylation, with the exception of a model built by us previously for eight ALL subtypes^30^. We compared the 519 CpG sites selected by ALLIUM to our previous classifier (n = 232 CpG sites), which resulted in 58.2% (135/232) overlapping sites with ALLIUM (Supplementary Table S22).

### Resolved molecular subtypes of B-other with ALLIUM

Next, we applied ALLIUM to the 150 remaining B-other cases in our cohort to investigate the distribution of recent ALL subtypes (Supplementary Table S23 and S24). For 64 cases where both DNAm and GEX data were available, 49 (76.6%) received concordant subtype predictions (**Figure 5a**). The highest concordance was observed for the prediction of subtypes with fusion genes, i.e. *ZNF384* (4/4, 100%), *KMT2A* (1/1, 100%), *DUX4*-r (11/12, 91.7%), *ETV6::RUNX1-like* (6/7, 85.7%), and PAX5alt (19/23, 82.6%). To establish consensus molecular subtypes for the B-other group, we constructed a 4-tier system to improve the confidence of subtype re-annotation (**Figure 5b**). Tier 1 included 29 patients with a high score from the DNAm or GEX classification combined with molecular evidence to support the subtype: expressed fusion gene, CNA, karyotype, or mutation. Tier 2 comprised 33 patients with concordant GEX and DNAm classification, but lacked conclusive molecular evidence. Tier 3 included 47 patients with only DNAm predictions or a discordant prediction with one non-class and one high score subtype prediction. Lastly, tier 4 included 41 patients where ALLIUM generated low confidence predictions or two conflicting predictions (**Figure 5c**).

**Figure 5.**
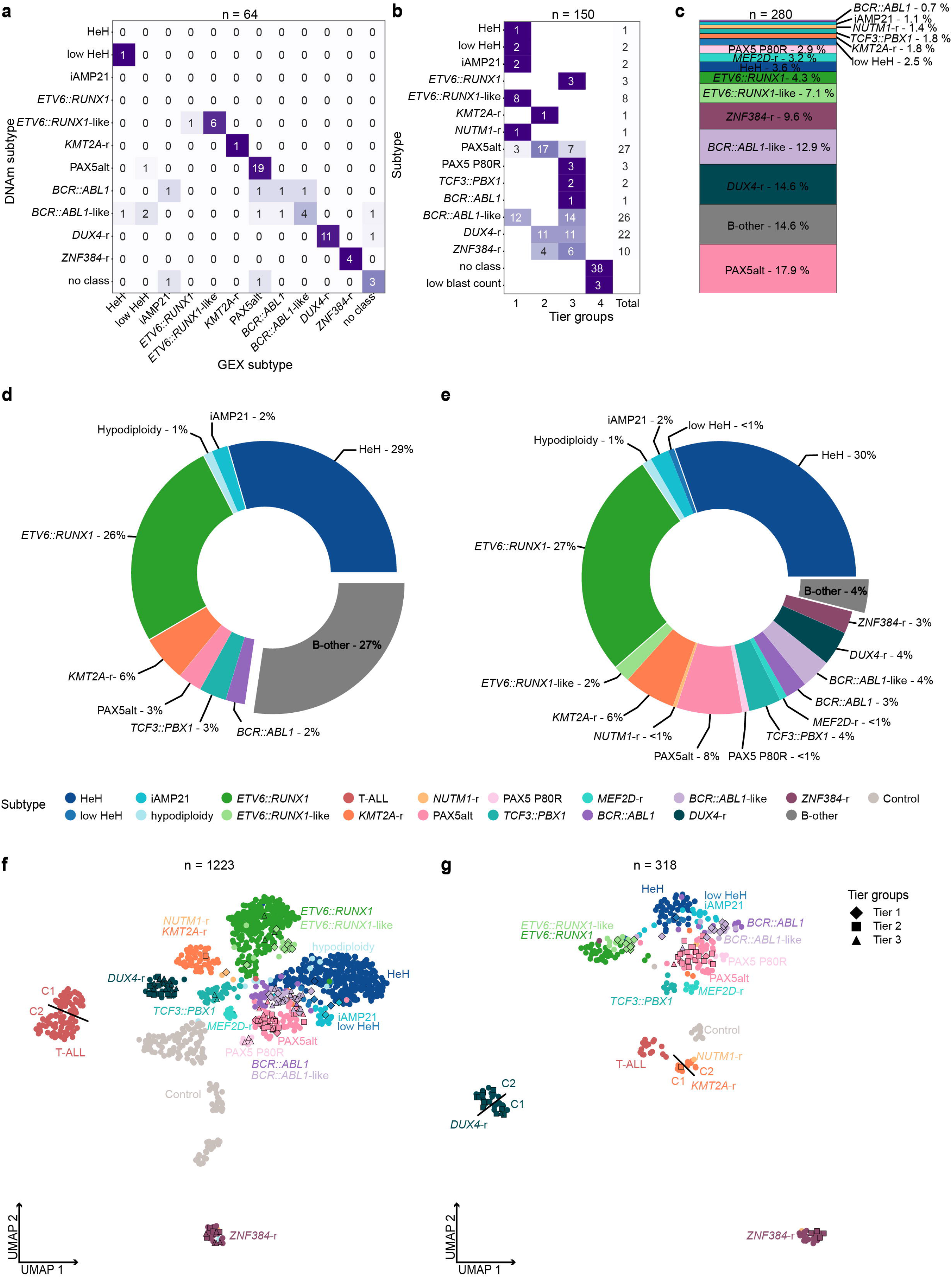
Frequencies of molecular subtypes. a) Molecular subtype concordance for 64 B-other patients based on DNA methylation (DNAm, x-axis) and gene expression (GEX, y-axis). b) Patient stratification into subtype and tier group for 150 B-other patients subtyped using ALLIUM. c) Distribution of all B-other patients (n = 280) after molecular re-classification. d) Subtype distribution of the BCP-ALL cohort (n = 1025) as was established at ALL diagnosis and e) after molecular re-classification. f) Unsupervised dimensionality reduction (UMAP) for samples with defined subtypes (n = 975), controls (n = 139) and newly re-characterized B-other patient samples (n = 109) based on DNAm levels of 519 CpG sites. g) UMAP based on 425 genes across samples with molecularly defined subtypes (n = 251), controls (n = 12) and newly re-characterized B-other patient samples by ALLIUM (n = 55).

In total, of the 280 B-other cases at the start of the study, 239 (85.4%) were assigned a new molecular subtype. Of these, 27 were found to harbor established subtype-defining aberrations prior to ALLIUM classification and 10 additional patients were identified after analysis with ALLIUM. Only 4% (41/1025) of our BCP-ALL cohort remained designated as B-other (**Figure 5d-e**).

The reclassified samples (tier 1–3) clustered with samples of known subtype (**Figure 5f-g**, Supplementary Figure S13). Sub-clusters were observed for *KMT2A*-r, *DUX4*-r, and T-ALL. In concordance with previous reports^46,47^, these included two putative *KMT2A*-r and *DUX4*-r subclusters based on GEX data and two T-ALL clusters in the DNAm data (**Figure 5f** and Supplementary Figure S13). Interestingly, the *KMT2A*-r and *DUX4*-r clusters were not visible in the DNAm visualization, while the T-ALL cluster was not visible in the GEX data, but this may be due to few (n = 19) patients with RNA-seq data. We examined fusion gene usage and found that the *KMT2A-r* cluster 1 (C1, n = 5) was characterized by *USP2* (n = 3) and *USP8* (n = 1) fusions, while C2 (n = 7) primarily contained patients with *KMT2A::AFF1* fusions (n = 4), indicating sub-clustering associated with the fusion partner. The patients in *DUX4*-r cluster 1 (C1, n = 12) expressed *DUX4-IGH* (n = 8), alongside a diverse array of other fusions, including *CRLF2-IRF1, PAX5-FLI1, ELL-KLF2, ATAD2-NPM1* and *PAX5-FOXP1*. In *DUX4*-r, cluster 2 (C2, n = 19) *DUX4-IGH* (n = 11) was the most prevalent fusions. Seven of the 19 T-ALL patients with RNA-seq data carried fusion genes, but no apparent clustering by fusion partner was observed.

### Outcomes of patients with new molecular subtypes treated on NOPHO protocols

Complete follow-up data was retrieved from 1125 out of the 1131 patients. The median follow-up time for the patients alive at the last follow-up was 16.0 years (IQR 13.0–19.0). The 5-year event-free survival (EFS) and overall survival (OS) of the entire cohort was 76.7% (95% CI 74.3%–79.2%) and 87.8% (95% CI 85.9%–89.7%), respectively (**Figure 6a**, Supplementary Table S25), with expected differences between NOPHO -92, -2000 and -2008 protocols (Supplementary Figure S14-S15). The 276 patients initially B-other at ALL diagnosis had significant differences in EFS and OS after stratification by new molecular subtype (p < 0.001, **Figure 6b**, Supplementary Table S25). Minimal residual disease (MRD) defined as > 0.1% at the end of induction (day 29) varied among the re-classified subtypes (Kruskal-Wallis p-value = 0.0004, **Figure 6c**). For example, only with PAX5alt did not have MRD (MRD+ 8.7%) yet this group had an intermediate EFS 77.6% (95% CI: 66.7%–90.2%) and OS 91.9% (95% CI: 84.5%– 99.8%). In contrast, 100% of *ZNF384*-r cases were MRD positive (n = 10) yet the EFS was 85.2% (95% CI: 72.8%–99.7%) and an excellent OS 96.3% (95% CI: 89.4%–100.0%) was observed. Survival analysis based on the two observed GEX *DUX4*-r sub-clusters (n = 31 patients, **Figure 5f)** showed differences in EFS between the groups (p = 0.048, Supplementary Figure S15). The most striking feature in this group was that three out of five patients who relapsed succumbed to their disease, while another three patients died in clinical remission (DCR1). Additional information can be found in the **Supplementary Tables and Figures** (Supplementary Table 25, Supplementary Figure S16).

**Figure 6.**
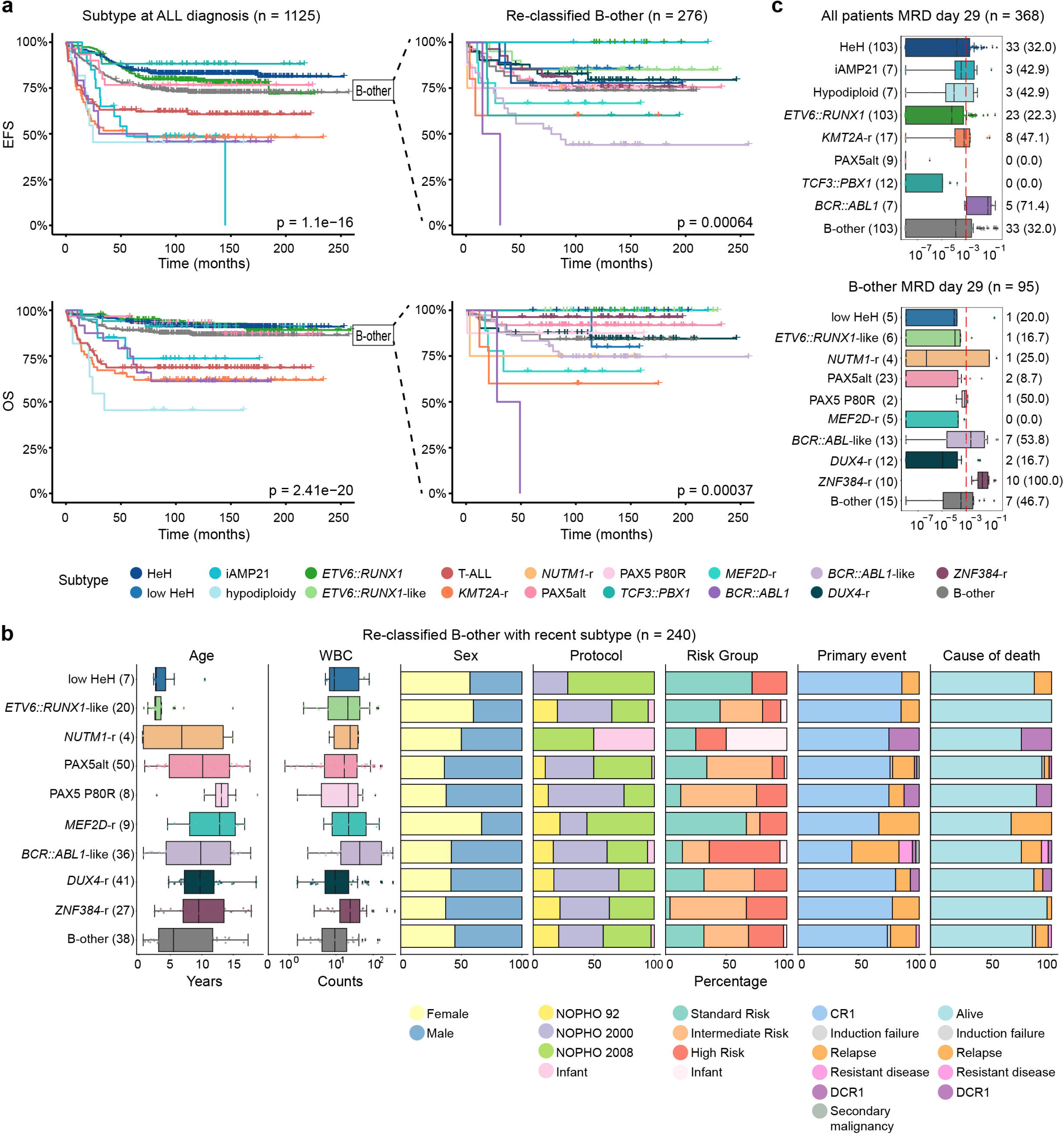
Clinical information and patient survival. a) Kaplan-Meir (KM) survival curves depicting event free survival (EFS, top) and overall survival (OS, bottom) colored according to molecular subtype as determined for 1125 patients at ALL diagnosis (left) and the re-classified B-other patients (n = 238) colored according to molecular subtype and the remaining unclassified “B-other” patients (n = 38, right). b) Clinical data including age, white blood cell count (WBC), sex, treatment protocol, risk groups, primary event, and cause of death per subtype for the B-other patients with recent subtype after re-classification (n = 240). CR1: complete remission, DCR1: death in complete remission. c) Minimal residual disease (MRD) at day 29 available for all patient data (n = 368, top) and re-classified B-other (n = 95, bottom) colored by their corresponding subtype.

## Discussion

Recent developments in integrated large-scale genomic analyses have greatly improved our knowledge of the genetic basis of ALL, identification of new subtypes and disrupted pathways that can be targeted therapeutically^20,24,46,48^. Accurate detection of the subtype-defining alterations in the clinical setting is crucial to guide risk and treatment stratification, monitor treatment response, and is very important for future implementation of tailored or precision therapy^2^. Given the low frequencies of rare subtypes in ALL and the long follow-up data needed to evaluate their clinical relevance, it is imperative to have methods that allow for retrospective analysis of biobank material, in addition to robust diagnostics in prospective cases. Herein, we designed and implemented a multimodal classification approach for ALL (ALLIUM) that captures epigenomic and transcriptomic alterations left as a detectable footprint in ALL cells. We demonstrate the utility of ALLIUM by retrospectively evaluating the frequency and clinical impact of emerging molecular cytogenetic subtypes in a large cohort of patients treated uniformly on NOPHO protocols between 1996 and 2013 and in external datasets.

Machine learning (ML) has potential to improve clinical diagnostics by enabling automated and accurate diagnostics, with reduced cost^49^. A unique feature of ALLIUM, over other ML-based subtype algorithms^26,27^ is that it can use multiple modalities (DNA methylation and/or gene expression) for subtype determination. We demonstrate here for the first time, that a DNAm-based classifier can achieve a comparable performance to GEX-based methods. A specific strength of DNAm as an analyte is its ability to identify disease-related methylation patterns and potential biomarkers in archived samples^33^. By using biobank samples and retrospective cohort studies, insight into long-term disease outcome can be gained, which would be difficult to obtain through prospective study designs, especially for rare subtypes. The ability of RNA-seq to detect fusion genes and coding mutations that can provide clear molecular evidence for subtype decision making, gives additional value to a GEX approach for prospective clinical diagnostics. However, RNA is not as readily available from historical material in biobanks, limiting the usefulness of GEX classifiers for retrospective interrogations. There is furthermore growing evidence to support methylation profiling for prognostication of T-ALL^47^, and array-based DNAm assays have the added benefit of generating CNA profiles^50^, which are helpful for diagnosing subtypes characterized by large-scale copy number changes, such as HeH, low HeH, hypodiploidy and iAMP21. In centers where DNAm subtyping for brain cancer is already routine^51,52^, ALLIUM DNAm subtyping could provide a complementary modality for routine disease diagnosis.

Using ALLIUM as a tool, we were able to accurately detect molecular ALL subtypes for up to 85.4% of previously unclassified (B-other) BCP-ALL cases in our population-based Nordic cohort spanning three NOPHO protocols (1992, 2000, 2008). We found that the molecular composition of BCP-ALL cases in the Nordics is comparable to studies from Europe^53,54^, USA^20,46,55^, and Asia^56^, and others^57^. In order of prevalence, these include PAX5alt (with a frequency of 8% compared to a range of 4–10% in the aforementioned studies), *BCR::ABL1*-like (4% vs 3–13%), *DUX4*-r (4% vs 4–7%), *ETV6::RUNX1*-like (2% vs 1–3%), *MEFD2*-r (<1% vs 1–2%), *NUTM1*-r (<1%, vs < 1–1%), PAX5 P80R (<1% vs 1–2%). Notably, we did not detect recently described very rare subtypes including t(5;14)(q31.1;q32.3)/*IL3::IGH, IKZF1* N159Y, or *CDX2-UBTF*^58^.

An additional strength of our study is our ability to assess the added value of MRD risk stratification in light of new molecular subtypes^37^. Although MRD remains as one of the best prognostic markers for treatment outcome in ALL, our results from patients treated on NOPHO-2008 further underscores that MRD stratification in the new subtypes may not be uniformly applicable^55,56,59^. Furthermore, early monocytic lineage switching, which includes loss of the B-cell immunophenotype, has been described in *DUX4*-r, *ZNF394*-r and *PAX5*-p80r subtypes^59^, potentially leading to an underestimation of MRD levels in these groups. However, questions still remain if MRD is a clinically relevant measure for treatment decisions in these new groups. Although we do not know what the MRD levels were of the patients included herein treated prior to 2008, our confirmatory observations further support that slow clearance of MRD specifically in the *ZNF384*-r may not accurately measure future outcome.

Excellent 5-year EFS and OS of outcomes of >95% have been reported for *DUX4*-r^46,55,56^. The lower 5-year OS (87.8%) for *DUX4*-r (n = 41) on the NOPHO protocols was unexpected, and will require additional investigation. Challenges also remain in identifying *DUX4* fusions due to the repetitive structure of the *DUX4* cassettes on chromosomes 4 and 10^12,54^. Using our modified read-mapping RNA-seq approach^9^, a *DUX4* fusion was detected only in ∼64% of cases. Several groups argue for a “*DUX4*-like” group in the absence of a detectible fusion gene, however we previously observed a case with a complicated insertion in between the *DUX4*-chr8q24.21-*IGH* transcript^9^, indicating that there are likely alternative formations of inserting a *DUX4* cassette into the IGH (or ERG) loci, which are not easily detected by short-read RNA-seq. Further investigation into these cases will be needed to improve the accuracy of diagnosing this important emerging subtype.

In summary, by implementing ALLIUM for retrospective analysis of a large retrospective ALL cohort treated uniformly on NOPHO protocols, we were able to accurately assess subtype distribution and long-term survival in new subtype groups. ALLIUM is freely available on GitHub and can be applied to determine molecular subtype membership of patients with either DNA methylation array data or RNA-seq data for research, or to support future precision diagnostics in pediatric ALL.

## Methods

### Patients

Bone marrow aspirates or peripheral blood samples collected at diagnosis from 1131 unique population.-based pediatric ALL patients were obtained from children diagnosed in the Nordic countries during 1996 –2013 and enrolled on the Nordic Society of Pediatric Hematology and Oncology (NOPHO) NOPHO-92 (n = 201), NOPHO-2000 (n = 493), NOPHO-2008 (n = 380), EsPh-ALL (n = 17), or Interfant (n = 40) treatment protocols^34–37^. Molecular diagnosis of ALL was established by analysis of leukemic cells at the time of diagnosis with respect to morphology, immunophenotype and cytogenetics. The guardians of the patients provided written or oral consent to the study. The study was approved by the regional ethics board in Uppsala, Sweden and by the NOPHO Scientific Committee (Study #56).

### DNA and RNA extraction

DNA and RNA were extracted from primary ALL cells after Ficoll gradient separation using reagents from the AllPrep DNA/RNA/miRNA Universal Kit (Qiagen) or the AllPrep DNA/RNA Kit (Qiagen) including a DNase treatment step (Qiagen). DNA and RNA were quantified using the reagents from the double stranded DNA Broad Range Kit or the RNA Broad Range kit on a Qubit instrument (Life Technologies). RNA quality was determined using the RNA Integrity Number (RIN) assessed by the Bioanalyzer or TapeStation system (Agilent).

### DNA methylation arrays

Genome-wide DNA methylation levels were determined using the Infinium HumMeth450K BeadChip assay (450k array, Illumina). DNAm data were generated using 250 ng input DNA from 384 newly collected BCP-ALL samples on the 450k array. Data from 741 patients were retrieved from Gene Expression Omnibus (GEO) entry GSE49031^45^. The complete DNAm dataset (1125 patients) was firstly filtered according to a previous study ^45^ resulting in 435,941 CpG sites and then to include probes present on the MethylationEPIC v.1.0. B5 manifest file (https://emea.support.illumina.com/downloads/infinium-methylationepic-v1-0-product-files.html), resulting in 406,542 CpG sites. Variance-based filtering removed 167,353 CpGs with low variability in the dataset (variance < 0.01). CNAs were detected using intensity levels from the 450k arrays using the R package “CopyNumber450kCancer”^60^. Data from 50 normal blood cell samples (CD3+ and CD19+, GSE49031) was used as control data for normalization and transformation of probe intensities (log2 ratio, LogR).

### RNA sequencing

A total of 328 samples from 315 patients were subjected to RNA sequencing (Supplementary Table S2 and S27). RNA sequencing libraries were prepared from 132 samples with RIN > 7 using the Illumina TruSeq stranded Total RNA (RiboZero human/mouse/rat) kit with 300 ng of total input RNA. The libraries were paired-end (PE) sequenced (150bp) on an Illumina HiSeq2500 or NovaSeq 6000 instrument to an average of 49.8M (range 30.2-113.2M) PE 150 bp reads per sample. Samples with RIN < 7 or with less than 300 ng input RNA available were prepared with the Illumina TruSeq RNA Access library preparation kit (n = 28 samples) and sequenced on an Illumina HiSeq 2500 instrument PE 150 bp to an average 34.0M (range 12.6-55.1M). RNA-seq data from 162 samples, generated with 1000 ng input RNA using the Script-Seq kit (EpiCentre)^9,38,61^ and 6 samples prepared with Illumina RNA access protocol^38,40^ were collected from previous studies. The raw sequencing data for each of the 328 libraries included in the study were processed together using the nextflow-based (21.02.0.edge) nf-core/rnaseq (3.0) pipeline, which includes trimming of the paired-end reads by trimgalore (0.6.6), alignment to GRCh38.103 with STAR (2.6.1d). The aligned reads were quantified at the transcript level using Salmon (1.4.0) and the transcript level expression values were subsequently summarized to the gene level using the bioconductor package tximeta (1.8.0). The gene count matrix was corrected for batch effects with ComBat-Seq. The genes were subsequently filtered to remove Y chromosome, scaffold, mitochondrial (MT), and ribosomal (RPS and RPL) genes, as well as non-protein coding genes resulting in 19,774 protein coding genes for downstream analysis. Data were normalized using Gene Length corrected trimmed mean of M-values (GeTMM), adjusting the data for both gene length and library size and finally log2 transformed. Technical (n = 5, repeated RNA-seq library construction from same RNA sample) and biological replicates (n = 8, sample taken at relapse) from 11 patients were used to validate merging the different library types (Supplementary Figure S17).

Fusion genes were detected using a combination of FusionCatcher 0.99.7d^62^ and targeted screening of 22 genes known ALL fusions (Supplementary Table S2). Fusion gene status for 61 patients in the study were described previously^7^. Candidate fusion genes were validated by supporting karyotype data, copy number analysis and/or by experimental validation using Sanger sequencing as previously described^9^.

### Mutational Analysis

Somatic single nucleotide variants (SNVs) were retrieved from a 872-cancer gene Haloplex panel for 144 patients in our study cohort^63,64^ and from whole genome sequencing performed on 41 patients^39,40,63^. Variant alleles *PAX5* p.Pro80Arg, *IKZF1* p.Asn159Tyr, and *ZEB2* p.His1038Arg were screened for in the 328 samples with RNA-seq data using alleleCount/3.2.2 (https://github.com/cancerit/alleleCount) on bam files.

### ALL subtype classification

ALLIUM was built using the scikit-learn package, based on the Nearest Shrunken Centroid (NSC) method^44,65^. Classifiers were built for each of 17 established molecular ALL subtypes present in our cohort and for healthy controls in a supervised manner. Models for DNAm and GEX datasets were designed separately. First, the data were split into design (known subtypes), hold-out (known subtypes) and discovery (B-other) sets. The models were trained, optimized and features were selected on the design set and then their performance was evaluated on hold-out and internal replication datasets. The models were further validated in independent external validation datasets: RNA-seq data from 65 Finnish patients for which detailed information can be found in the Supplementary Materials and Methods, and published datasets from RNA-seq of 19 BCP-ALL patients from GSE161501^42^ and 450k DNAm from 227 BCP-ALL patients GSE56600^43^. Additional details can be found in the **Supplementary Materials and Methods**. ALLSorts and ALLCatchR were run on the corrected count matrix (n genes = 60,666) according to the instructions (https://github.com/Oshlack/ALLSorts/wiki/1.-Installation, https://github.com/ThomasBeder/ALLCatchR)^26,27^.

### Survival analysis

Outcome data for the 1131 patients in the study was retrieved from the NOPHO leukemia database in February 2022. In total, 1125 patients had complete follow-up data available, the average time since diagnosis was 16.3 years (range 9-26). OS was calculated as the time from the date of diagnosis to the date of last follow-up or death of any cause. EFS was calculated from the date of diagnosis until an event (relapse, induction failure, resistant disease, secondary malignant neoplasm) as defined in the protocol in question, death during induction, or death during complete remission or the date of last follow-up for patients without event. Patients who received allogeneic stem cell transplant (allo-SCT) were not censored at the time of transplant. OS and EFS curves were generated using the Kaplan–Meier estimation, and the log-rank test was used for comparisons between groups. Survival analysis was performed in R using the survival and survminer packages. Kruskal-Wallis H-test from the Python library scipy.stats assessed the significance of subtype-stratified MRD distributions. A p-value <0.05 (2-tailed) was considered statistically significant.

## Supporting information

Supplementary Methods and Results

## Data Availability

All data produced in the present study are available upon reasonable request to the authors.

## Data availability

The 450k DNA methylation data are available under controlled access via 10.17044/scilifelab.22303531. The GEX count matrix is available in GEO under the accession number GSE227832. Requests for data sharing may be submitted to Jessica Nordlund.

## Code availability

All scripts and environment requirements to reproduce the analyses, as well as the ALLIUM model are available at GitHub https://github.com/Molmed/Krali_2023.

## Acknowledgements

This work was supported by grants from the Swedish Research Council (2019-01976 to JN), the Swedish Cancer Society (CAN2018-623 to ACS and CAN2022-2395 to JN), the Swedish Childhood Cancer Foundation (PR2017-0023 to ACS and PR2019-0046 to JN), the Göran Gustafsons Foundation (to JN), the Jane and Aatos Erkko Foundation and the Academy of Finland #321550 (to OL and MH). DNA methylation array analysis and RNA-sequencing was performed with assistance from the SciLifeLab National Genomics Infrastructure, SNP&SEQ Technology Platform, which is funded by the Swedish Research Council and the Knut and Alice Wallenberg Foundation. Computational resources were provided by the Swedish National Infrastructure for Computing (SNIC) and the Finnish IT Centre of Science (CSC) and University of Eastern Finland Bioinformatics Center. We thank Sara Nystedt and Sara Nilsson for technical assistance, Jonas Carlsson Almlöf and Christofer Bäcklin for input on DNA methylation classification, and our colleagues from NOPHO LL Biology Group, Lucia Cavelier, Anna Bremer and Tatjana Pandzic for valuable input on the study design. We especially thank the ALL patients who contributed samples to this study. Figure 1 and Supplementary Figures S2-S3 were made with Biorender.com.

## Author information

### Contributions

YMZ, GL, ACS, and JN conceived the study. OK, YMZ, GA, APE, AL, SS, VZ, IIÖ, and LO analyzed the data. OK designed ALLIUM and made the figures. YMZ, AL, VZ, LO, and KV performed experiments. VZ, MH, JS, IIÖ, GB, AN, LO, KV, HL, TFi, and OL provided RNA-seq data. VZ, IIÖ, GB, AN, and UNN provided expertise on clinical diagnostics. HOM and HVM provided MRD data. TFl, EF, OGJ, JK, OL, UNN, KS, AH, MH, and GL provided clinical material and data. MH provided information from the NOPHO registry and assisted with survival analysis. EF and UNN provided karyotyping expertise and data. MH, OL, GL, ACS, and JN secured funding. OK, YMZ, APE, GA, GL, and JN wrote the manuscript. All authors read and approved the final version.

## Ethics declarations

### Competing interests

The authors declare no competing interests.

## Notes

### Competing Interest Statement

The authors have declared no competing interest.

### Funding Statement

This study was funded by the Swedish Research Council (2019-01976), the Swedish Cancer Society (CAN2018-623 and CAN2022-2395), the Swedish Childhood Cancer Foundation (PR2017-0023 and PR2019-0046), the Göran Gustafsons Foundation, the Jane and Aatos Erkko Foundation and the Academy of Finland (#321550).

### Author Declarations

The study was approved by the regional ethics board in Uppsala, Sweden and by the Nordic Society of Paediatric Haematology and Oncology (NOPHO) Scientific Committee (Study #56).

