## Supplementary Figures for "Multimodal classification of molecular subtypes in pediatric acute lymphoblastic leukemia"

*Corresponding author:

Dr. Jessica Nordlund

Box 1432, BMC

75144 Uppsala, Sweden

### Supplementary Figures

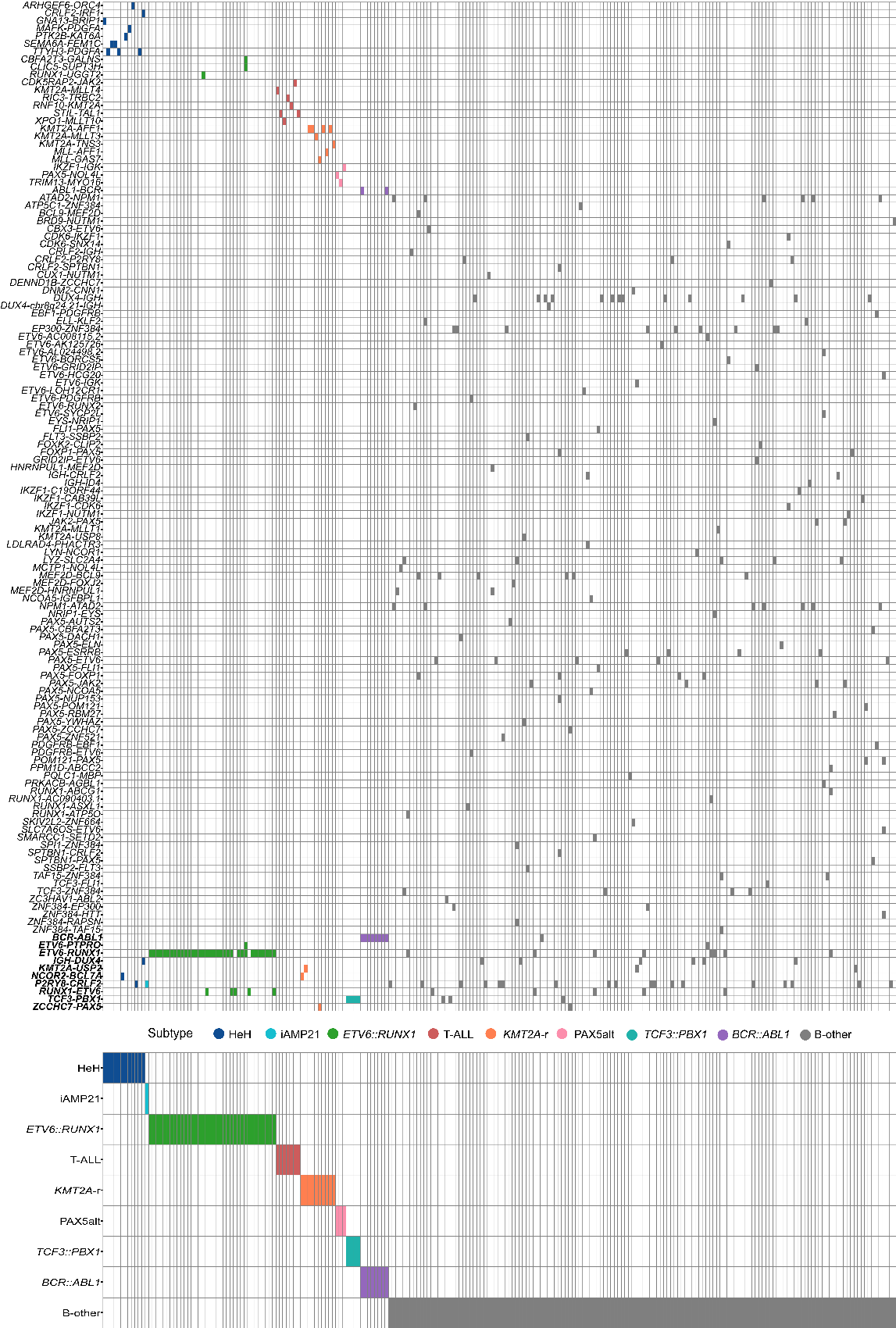

**S1. Fusion gene distribution across 225 ALL patients labeled by subtype received at ALL diagnosis. The patients are denoted in columns and fusion genes in rows.** In total 131 unique fusion genes (including the reciprocal cases) were detected. Ten fusion genes appeared across multiple subtypes (denoted with bold Italics on the bottom).

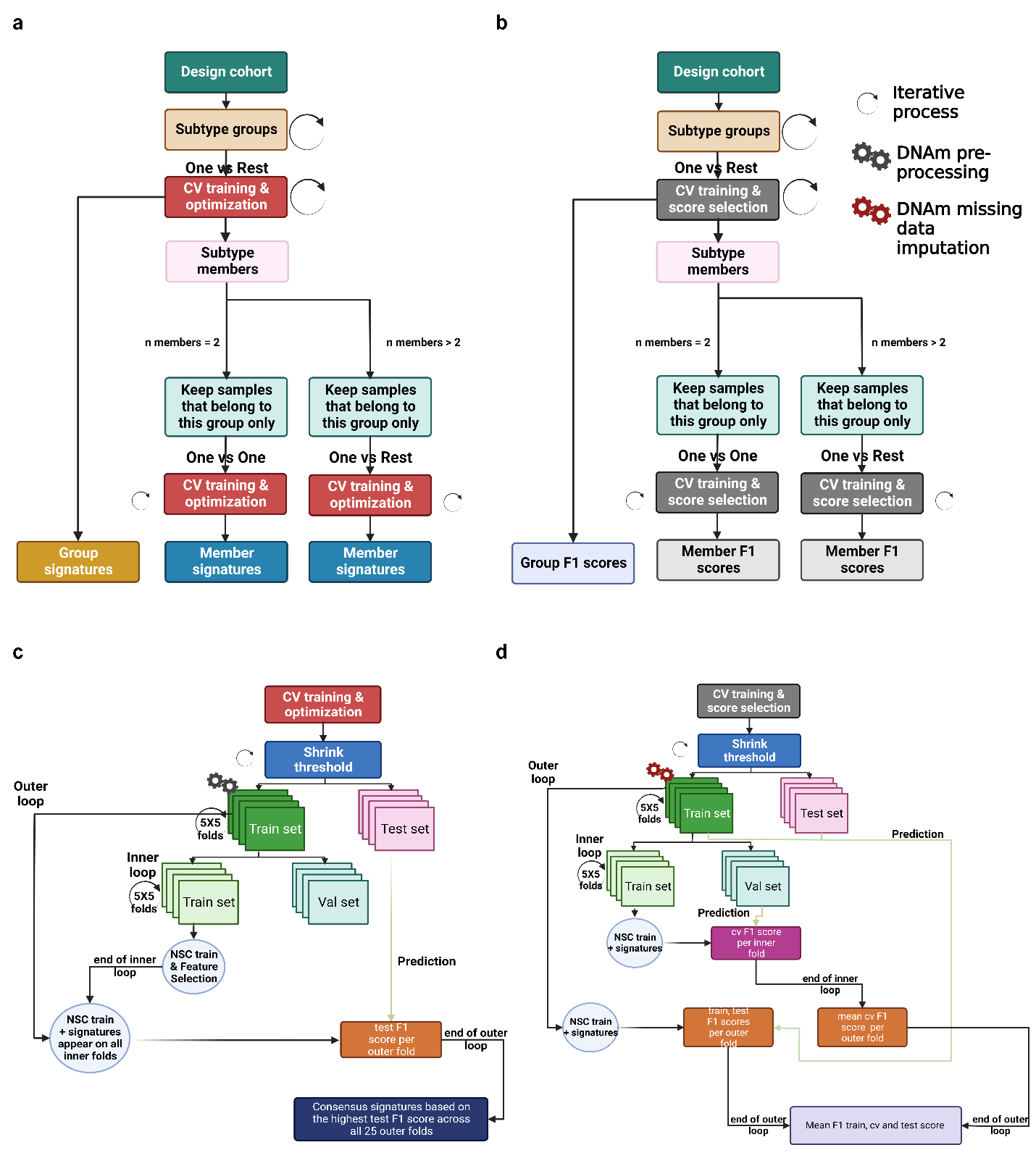

**S2. ALLIUM classifier optimization and feature selection.** a) Overview of cross validation (CV) and optimization iterative steps. b) Overview of CV and F1 score selection iterative steps. (a) and (b) utilize a multi-step approach for groups of subtypes with similar biology and subtype members or single subtype groups to obtain group and member signatures. c) During optimization, an iterative CV approach captures signatures for each shrink threshold parameter per group/subtype. d) External CV to obtain the train, test and CV F1 scores per shrink threshold based on the selected signatures from (c) (Created with BioRender.com).

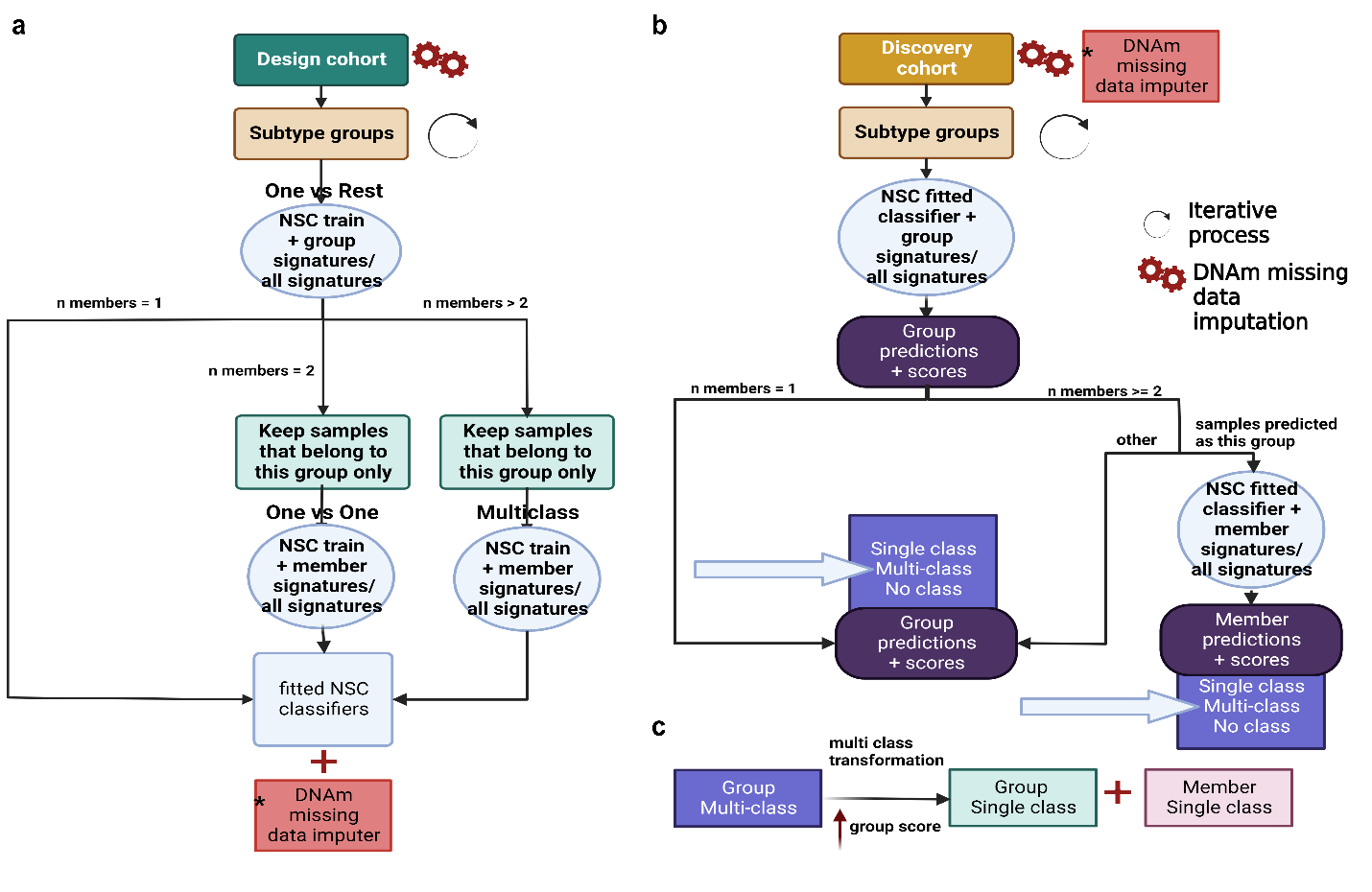
**S3. Final ALLIUM classifier model architecture.** a) ALLIUM was trained on all design cohort samples in a multi-step procedure including a group and a subtype membership step. When the train mode is set to 'all', all selected signatures are used to train each classifier, otherwise each classifier uses its own signatures. b) Subtype prediction is based on the trained models outlined in panel (a). c) The multi-class to single class transformation assigns subtype based on the highest group probability score. (Created with BioRender.com).

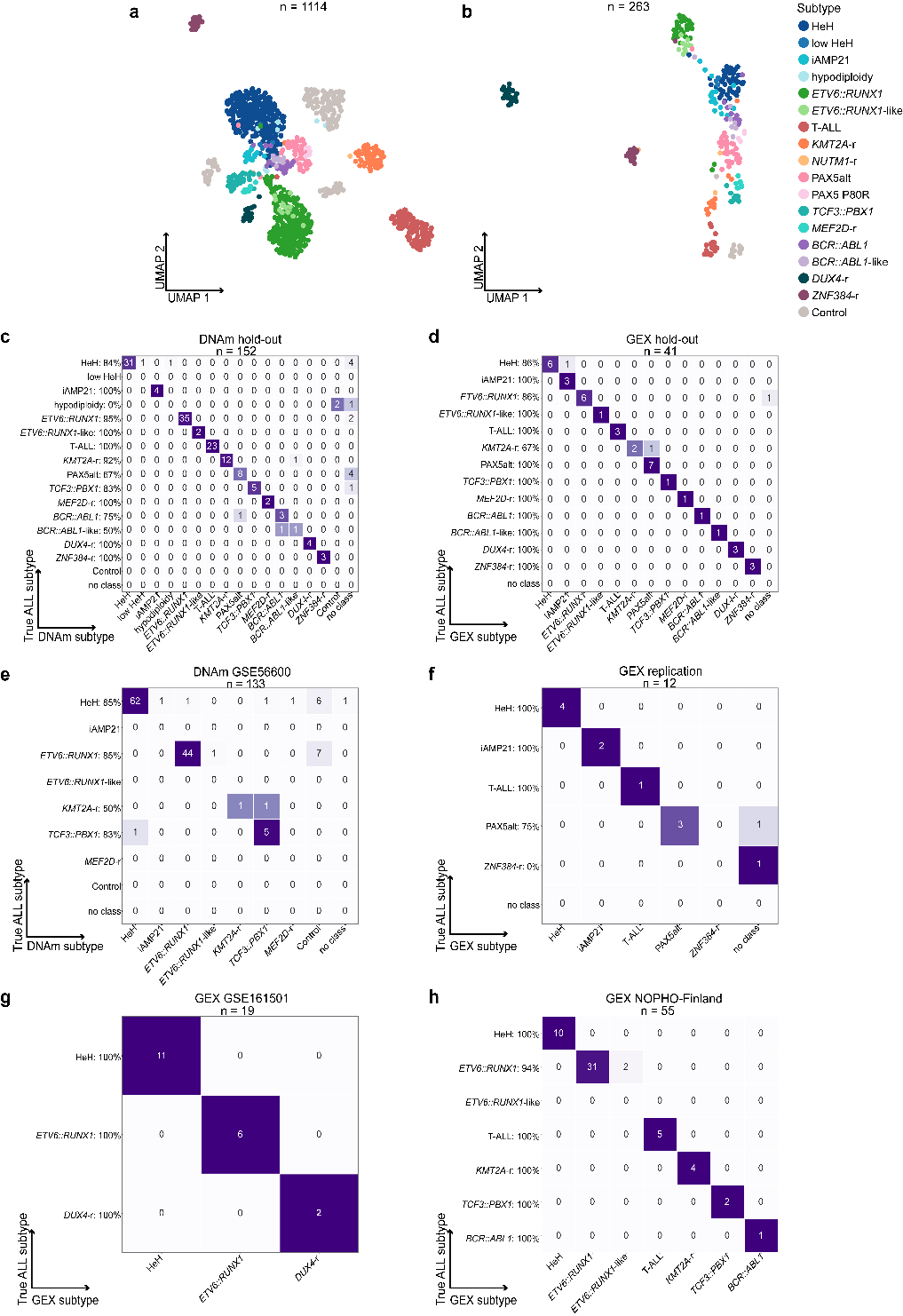

**S4. ALLIUM performance evaluation.** a) Unsupervised dimensionality reduction (UMAP) based on DNA methylation (DNAm) levels of 519 CpG sites across molecularly defined patients (n = 975) and controls (n = 139). b) UMAP based on gene expression (GEX) levels of 425 genes across molecularly defined patients (n = 251) and controls (n = 12). c) DNAm vs true cytogenetic subtypes for the hold-out dataset (87.5% concordance, 133/152). d) GEX predictions vs true cytogenetic subtypes for the hold-out dataset (92.7% concordance, 38/41). e) DNAm predictions vs true cytogenetic subtypes for the DNAm GSE56600 dataset (84.2 % concordance, 112/133). f) GEX predictions vs true cytogenetic subtypes for the internal replication dataset (83.3% concordance, 10/12). g) GEX predictions vs true cytogenetic subtypes for the GEX GSE161501 dataset (100%, 19/19). h) GEX predictions vs true cytogenetic subtypes for the GEX NOPHO-Finland dataset (96.4%, 53/55).

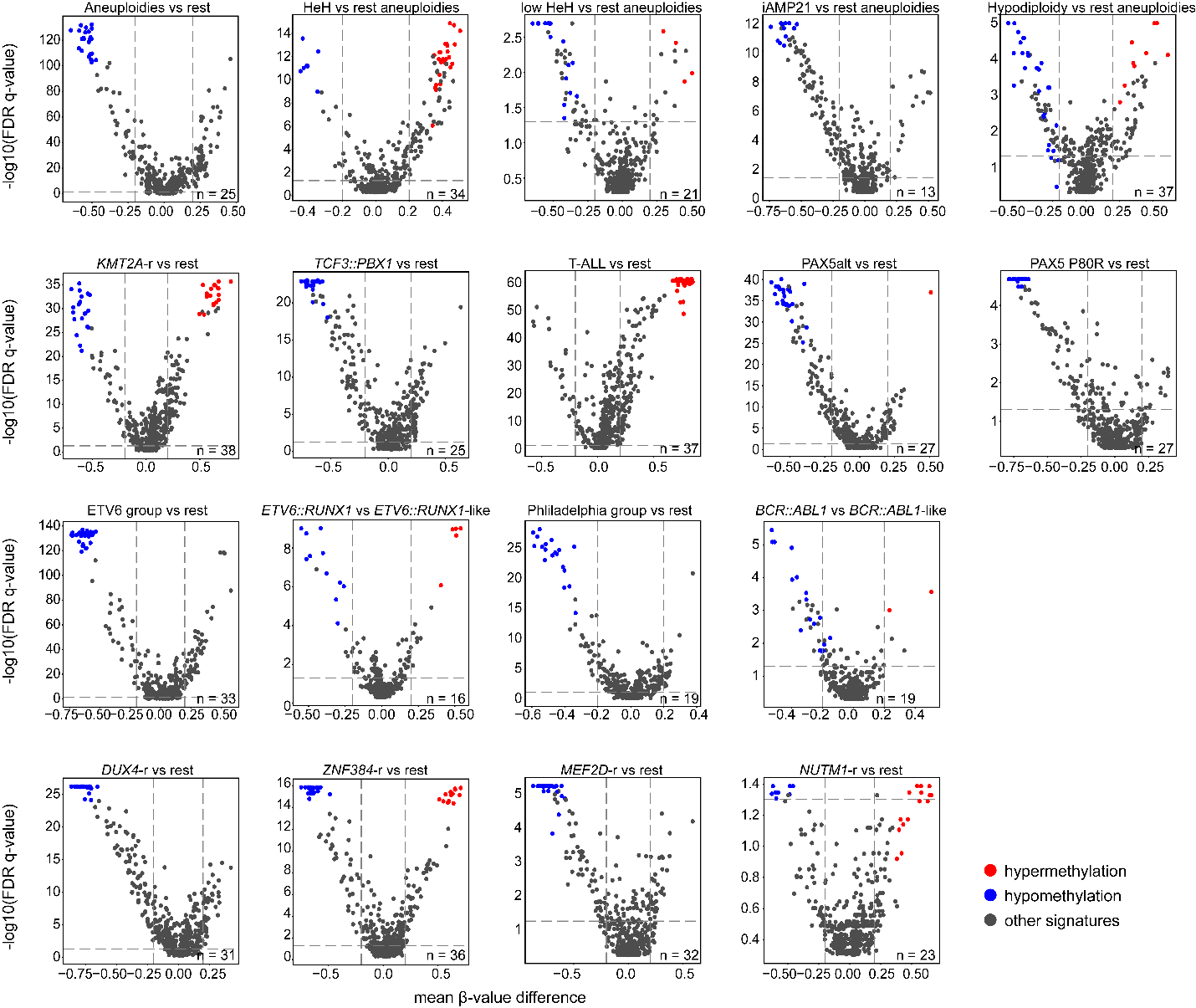

**S5. Volcano plots for the CpG sites selected by ALLIUM.** Mean methylation β-value difference (x-axis) is plotted against the Benjamini-Hochberg (BH) corrected Mann-Whitney U test p-value (y-axis) for the 1084 patients with established molecular subtypes after ALLIUM analysis across the 493 subtype-defining CpG sites (excl. control CpGs, n = 26). Each panel is labeled by the groups or subtypes compared and the total number of CpGs selected by ALLIUM are shown at the right bottom of the volcano plot. The CpG sites are colored according to the key to the right of the figure, where sites with β-value difference > 0 are colored in red (hypermethylated) and those < 0 are colored in blue (hypomethylated). The remaining CpG sites that were not selected to differentiate the subtype plotted are colored in grey. Signatures meeting the adjusted p-value threshold of < 0.05 and absolute β-value difference > 0.2 are located on the upper right (significantly hypermethylated) or upper left (significantly hypomethylated) rectangles defined by the threshold dashed lines.

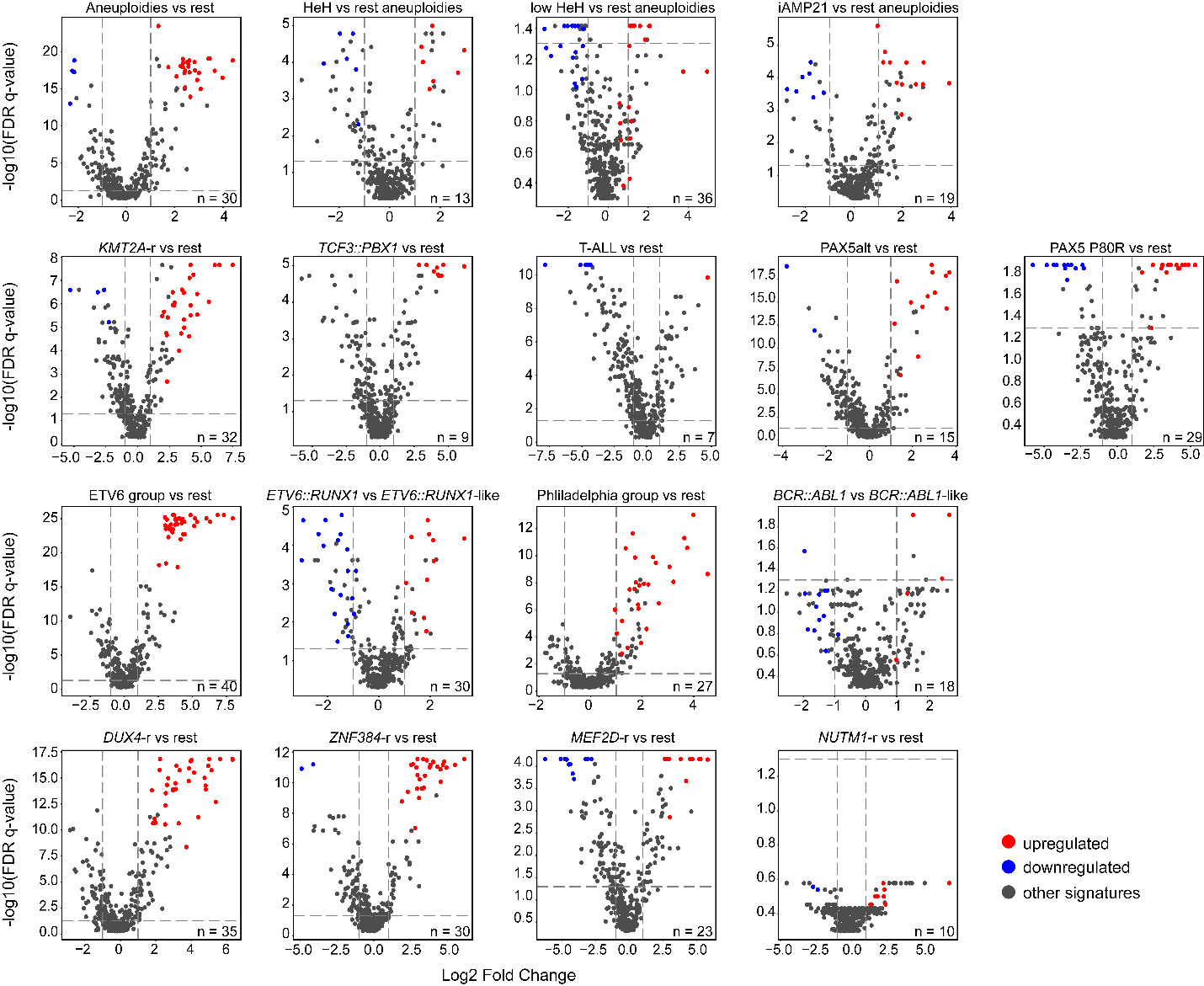

**S6. Volcano plots for expressed genes determined by ALLIUM.** Mean log2 fold change (log2FC, x-axis) is plotted against the Benjamini-Hochberg (BH) corrected Mann-Whitney U test p-value (y-axis) for the 306 patients with established molecular subtypes after ALLIUM analysis across the 403 subtype-defining genes high-lighted by the GEX classifier (excl. control CpGs, n = 22). Each panel is labeled by the groups or subtypes compared. The genes colored according to the key to the right of the figure, where genes with log2 change > 0 are colored in red (upregulated) and log2 change <0 in blue (downregulated). The remaining genes that were not selected for the subtype of interest are colored in grey. Signatures meeting the adjusted p-value threshold of < 0.05 and absolute log2 foldchange > 1 are located on the upper right (significantly upregulated) or upper left (significantly downregulated) rectangles defined by the threshold dashed lines.

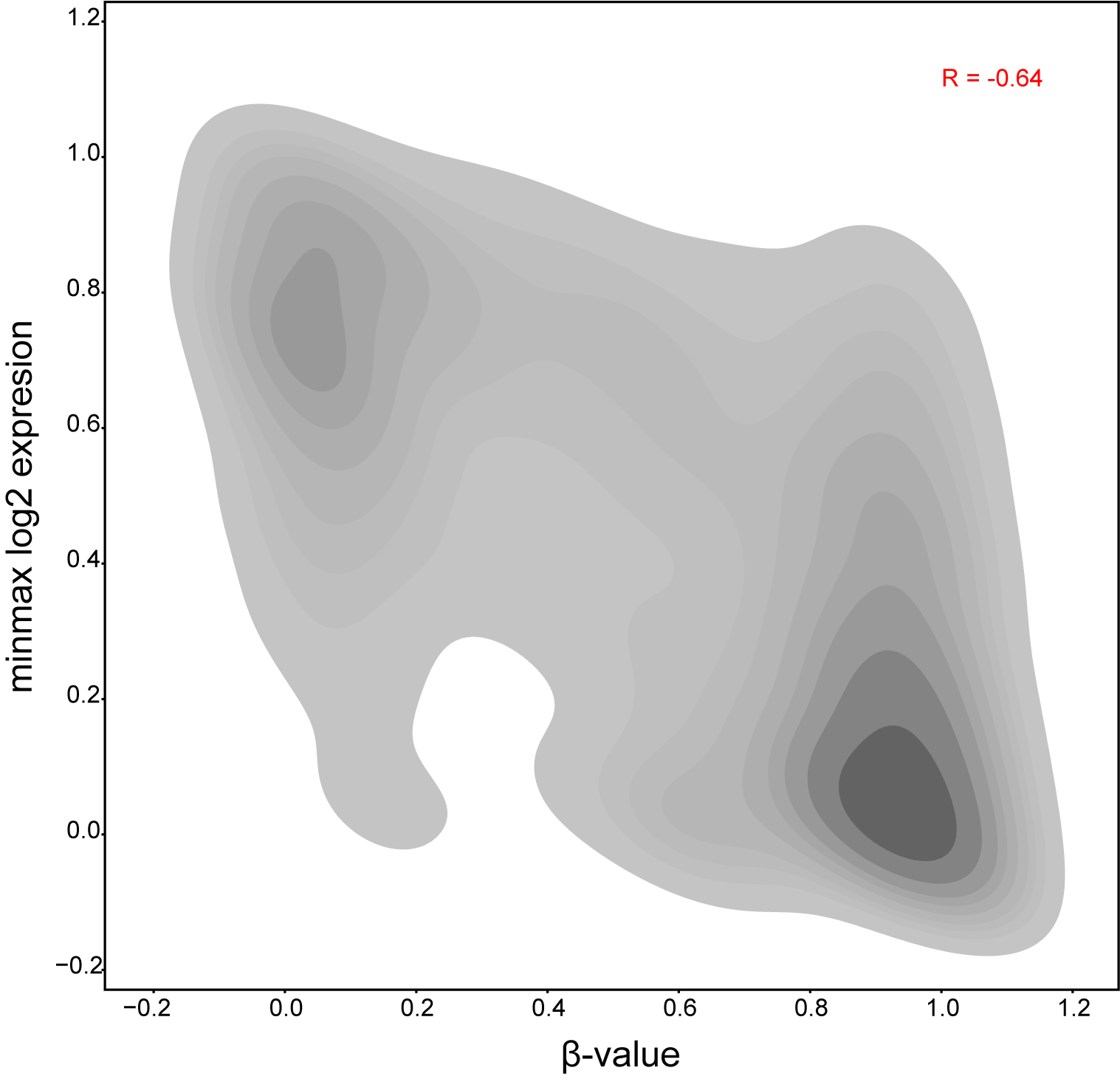

**S7. Correlation between DNAm and GEX.** The bivariate kernel density estimate (KDE) plot depicts the correlation of DNAm vs GEX by visualizing their distribution (β-value (x-axis) and GEX minmax scaled log2 expression (y-axis) per patient gene-CpG pair). **The beta-values for 22 CpGs were compared to the GEX levels of their closest mapping gene (12 genes in total) selected by ALLIUM DNAm and GEX classifiers for 204 patients with DNAm and GEX data available.** Pearson’s correlation coefficient was utilized to calculate the overall correlation between DNAm and GEX.

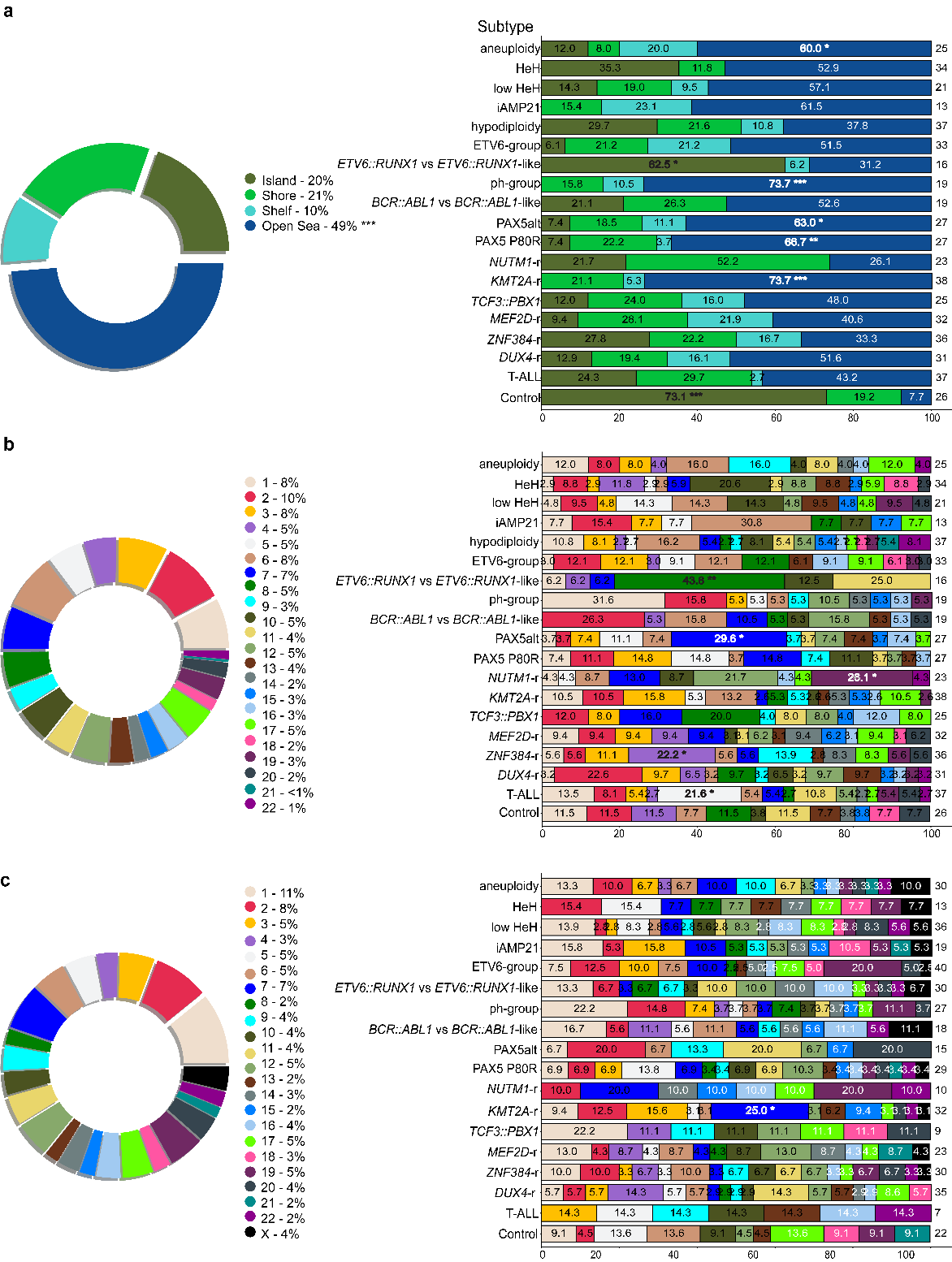

**S8. Annotation of ALLIUM DNAm and GEX features.** a) Vicinity to CpG islands of the 519 ALLIUM DNAm CpG sites in percentage across all subtypes (left) and per signature group (right). b) The chromosomal location of the 519 CpG sites in percentage (left) and per signature group (right). c) The chromosomal location of the 425 ALLIUM GEX genes in percentage (left) and per signature group (right). The number of signatures per group is denoted on the right of each stacked bar plot. Significantly enriched cases compared to the proportions prior signature selection by ALLIUM classifiers are denoted with asterisk(s) (FDR p-value: *** < 0.001, ** < 0.01, * <0.05).

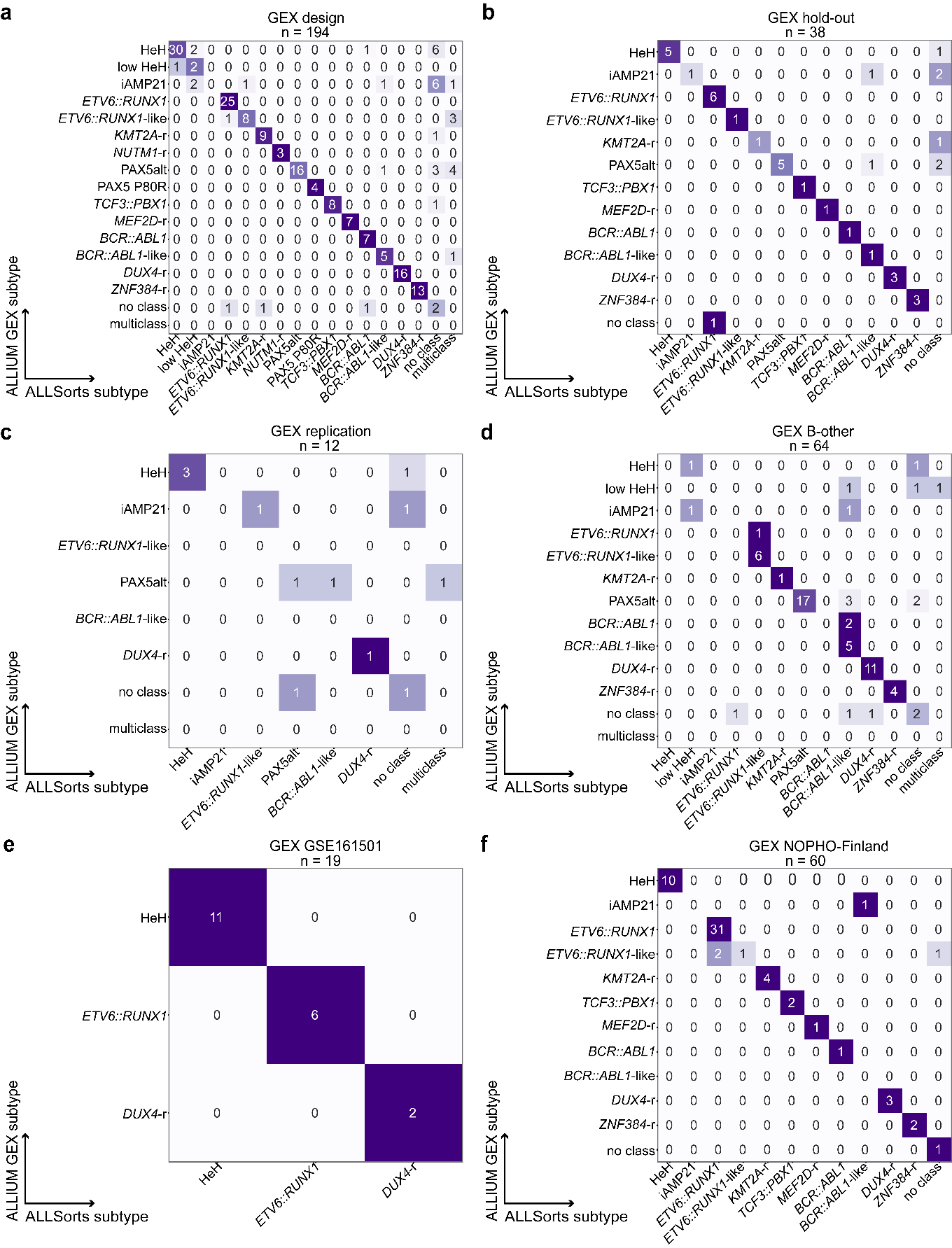

**S9. GEX predictions by ALLIUM and ALLSorts**. a) Design dataset 79.9% concordance (155/194). b) Hold-out dataset 76.3% concordance (29/38). c) Replication dataset 50% concordance (6/12). d) Discovery (B-other) dataset 71.9% concordance (46/64). e) GEX GSE161501 100% concordance (19/19). f) GEX NOPHO-Finland dataset 93.3% concordance (56/60). T-ALL patients (n = 25) were excluded.

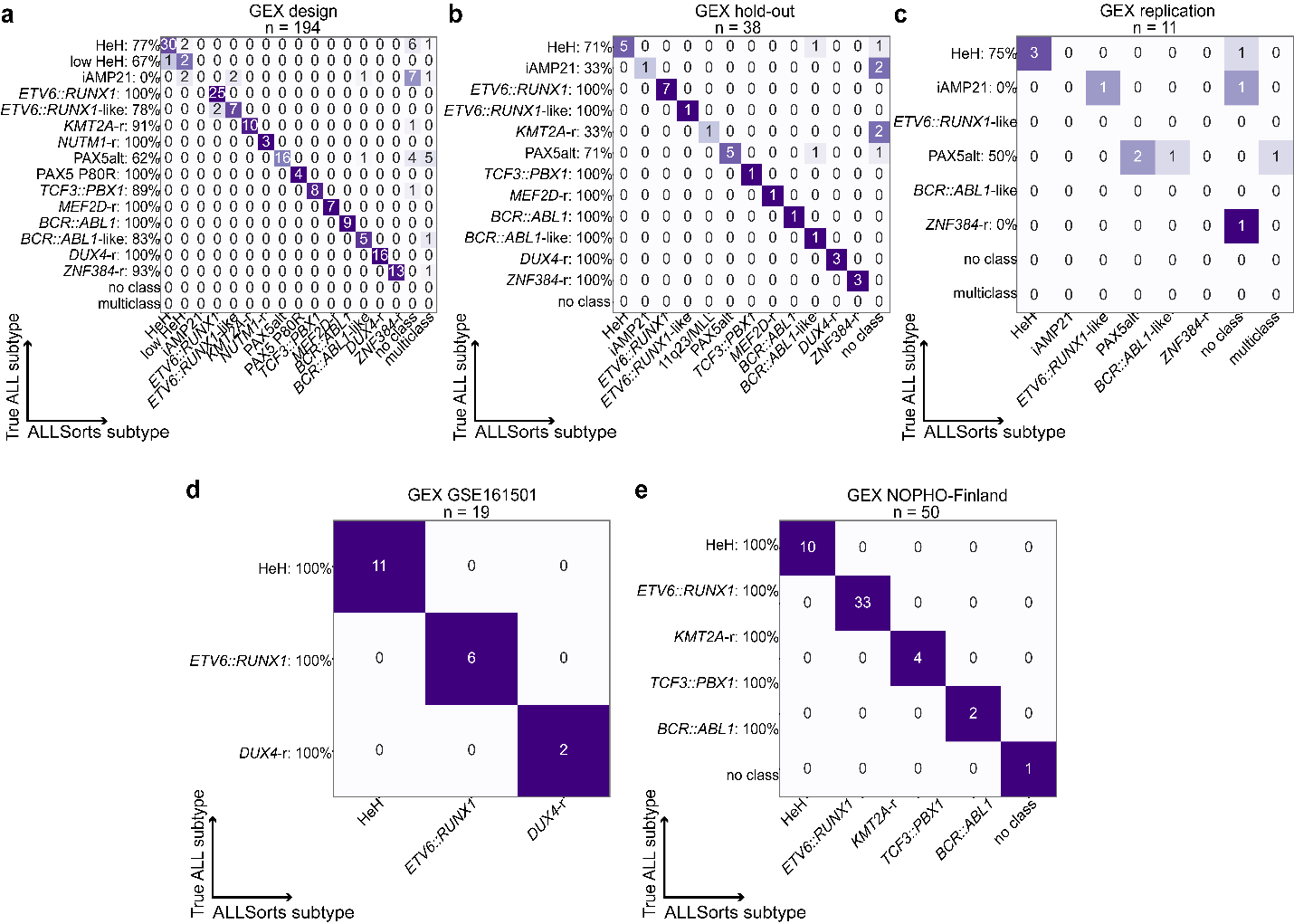

**S10. GEX predictions by ALLSorts vs true cytogenetic subtype.** a) Design dataset (79.9% concordance, 155/194). b) Hold-out dataset (78.9% concordance, 30/38). c) Replication dataset (45.5% concordance, 5/11). d) GEX GSE161501 dataset (100% concordance, 19/19). e) GEX NOPHO-Finland dataset (100.0% concordance, 50/50). T-ALL patients (n = 25) were excluded.

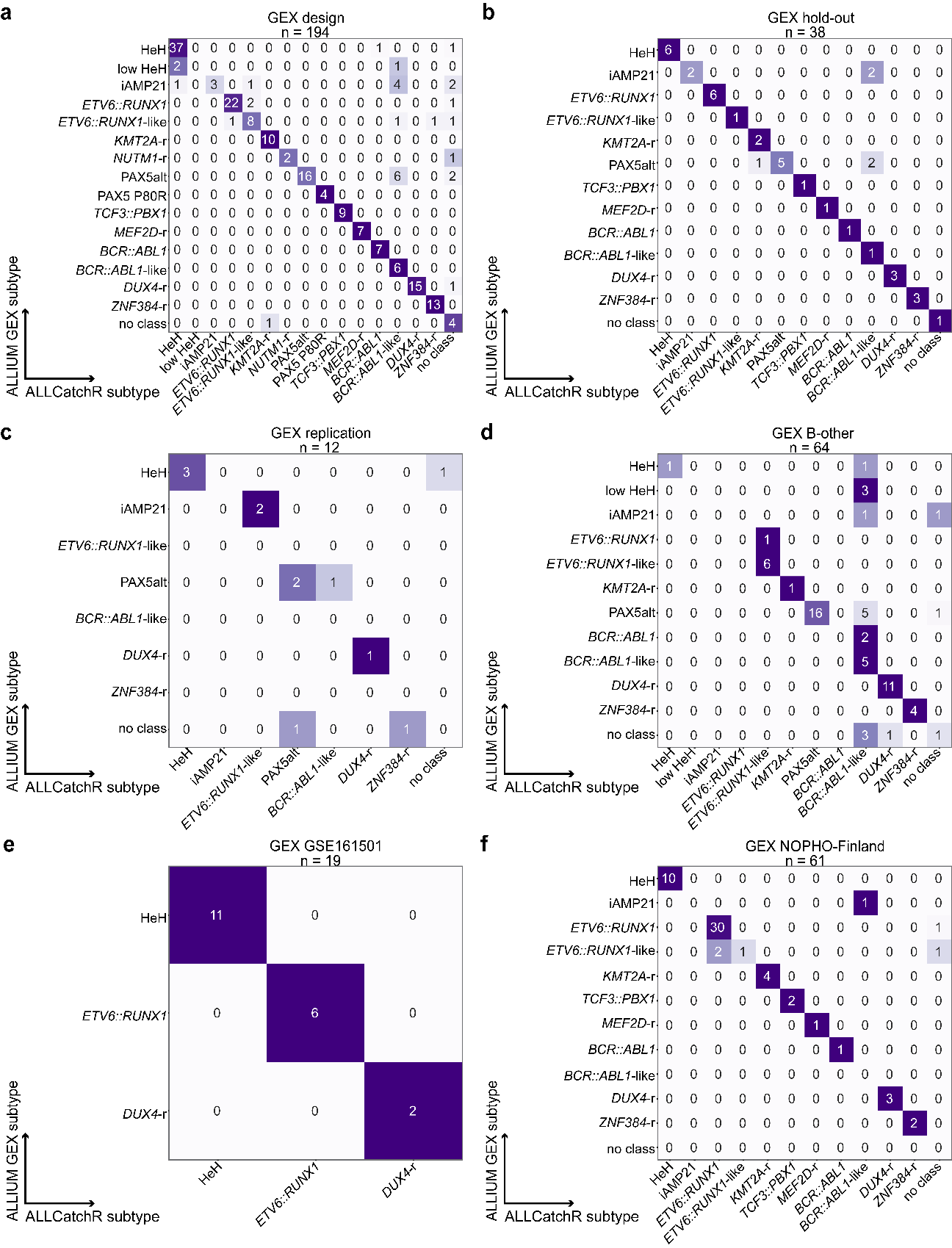

**S11. GEX predictions for ALLIUM and ALLCatchR**. a) Design dataset 84.0% concordance (163/194). b) Hold-out dataset 86.8% concordance (33/38). c) Replication dataset 50% concordance (6/12). d) Discovery (B-other) dataset 70.3% concordance (45/64). e) GEX GSE161501 100% concordance (19/19), f) GEX NOPHO-Finland dataset 90.0 % concordance (54/60). T-ALL patients (n = 25) were excluded.

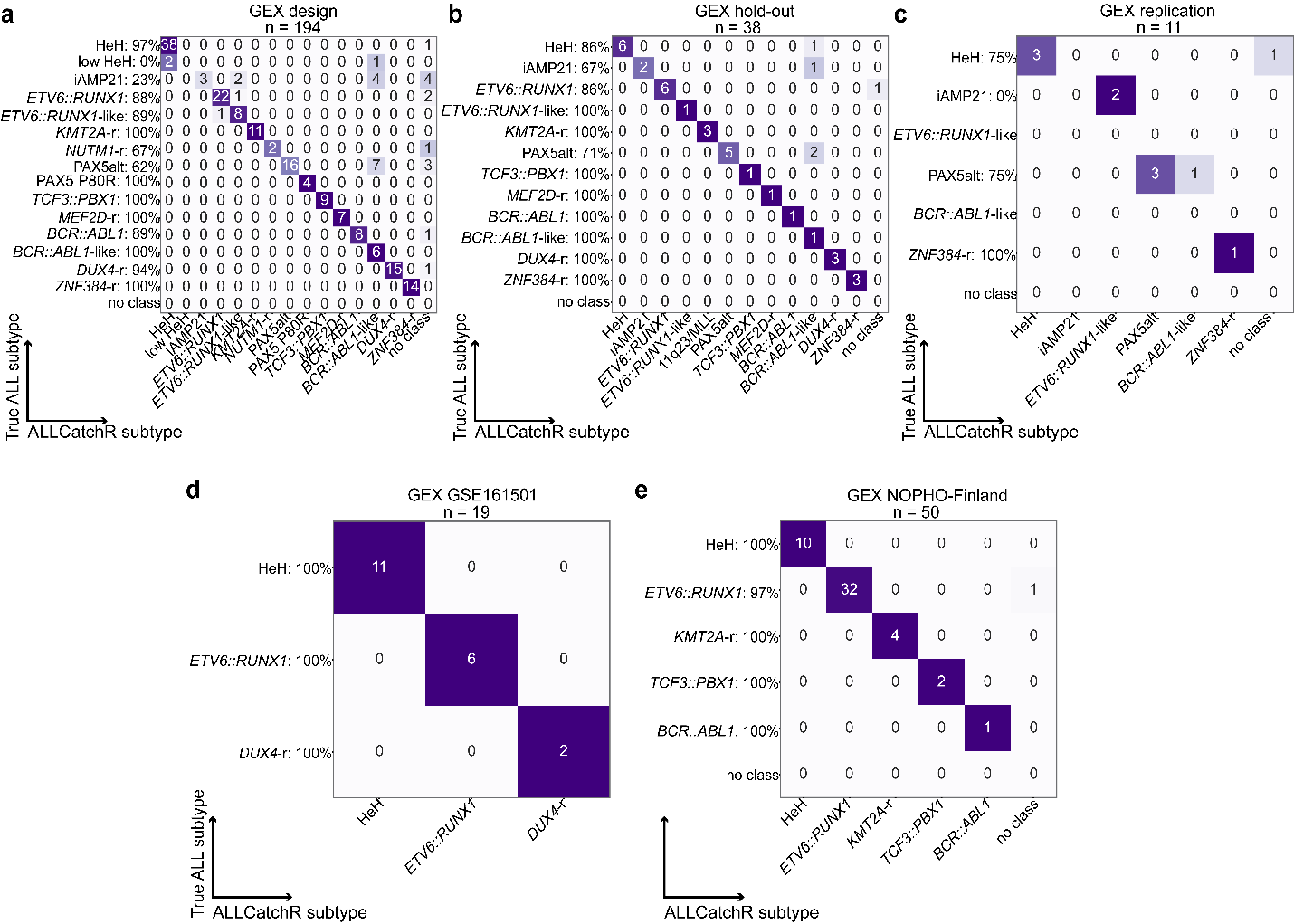

**S12. GEX predictions by ALLCatchR vs true cytogenetic subtype.** a) Design dataset (84.0% concordance, 163/194). b) Hold-out dataset (86.8% concordance, 33/38). c) Replication dataset (63.6% concordance, 7/11). e) GEX GSE161501 dataset (100%, 19/19). f) GEX NOPHO-Finland dataset (98.0%, 49/50). T-ALL patients (n = 25) were excluded.

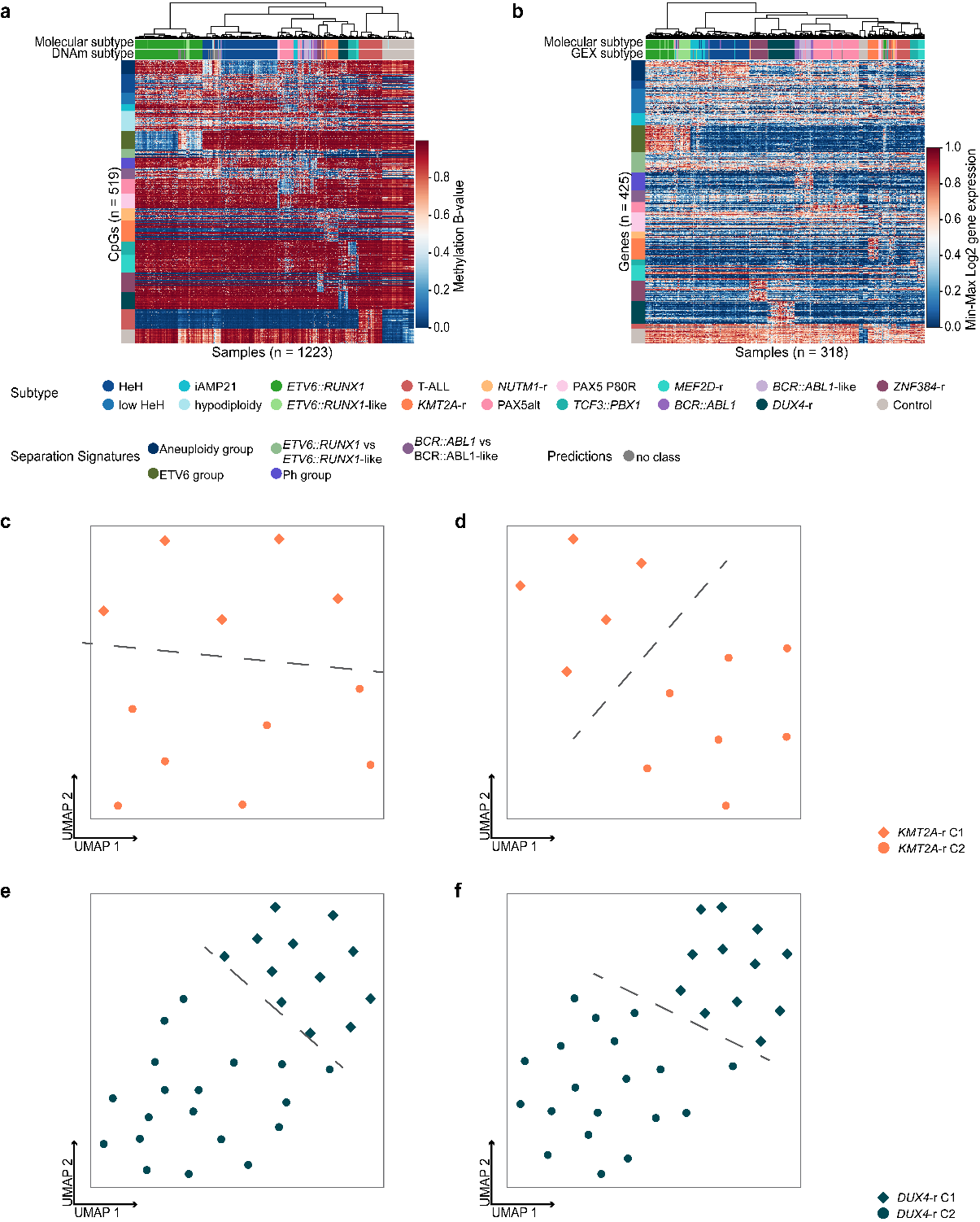

**S13. Unsupervised visualization after re-classification of B-other.** a) Unsupervised hierarchical clustering based on DNAm levels of 519 CpG sites across molecularly defined (n = 975), control (n = 139) and newly re-characterized B-other samples (n = 109). b) Unsupervised hierarchical clustering based on GEX levels of 425 genes across molecularly defined (n = 251), control (n = 12) and newly re-characterized B-other samples (n = 55). c) UMAP plot based on the 425 ALLIUM GEX genes and 12 patients with *KMT2A*-r. Two clusters C1 (n = 5) and C2 (n = 7) are highlighted. d) UMAP plot based on 2,695 genes for distinguishing *KMT2A*-r clusters as determined by Brady et al^1^. The same two clusters are observed in panels C and D. e) UMAP plot based on the 425 ALLIUM genes across 31 *DUX4*-r samples. Two clusters are observed, C1 (n = 12) and C2 (n = 19). f) UMAP plot based on the 2,039 genes used for distinguishing *DUX4*-r clusters as determined by Brady et al^1^.

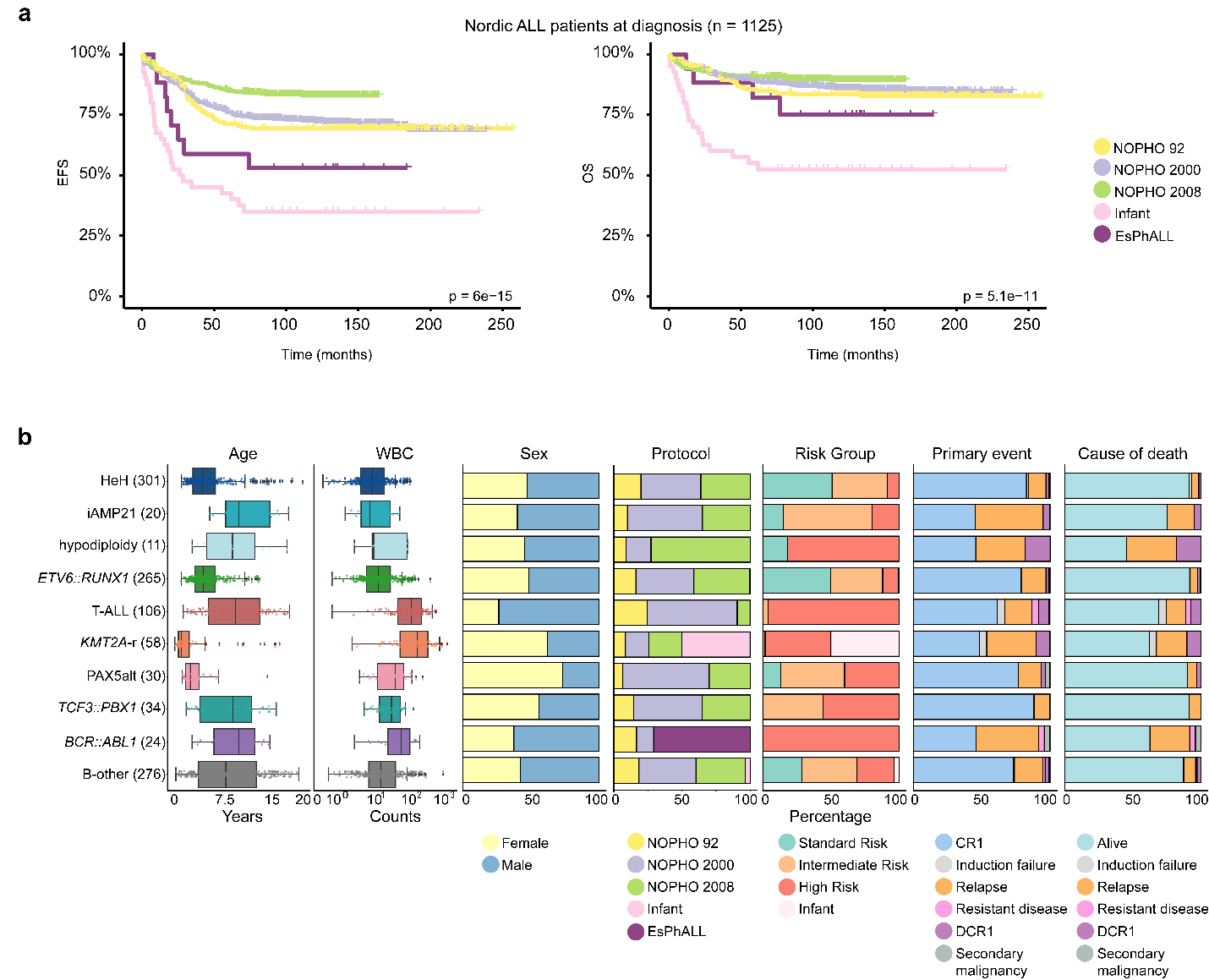

**S14. 1125 patients treated according to NOPHO protocols.** a) Kaplan-Meier survival curves depicting Event Free Survival (left) and Overall Survival (right) stratified by treatment protocol. b) Clinical data including age, white blood cell count (WBC), sex, treatment protocol, risk groups, relapse, and cause of death for the 1125 patients with complete survival data.

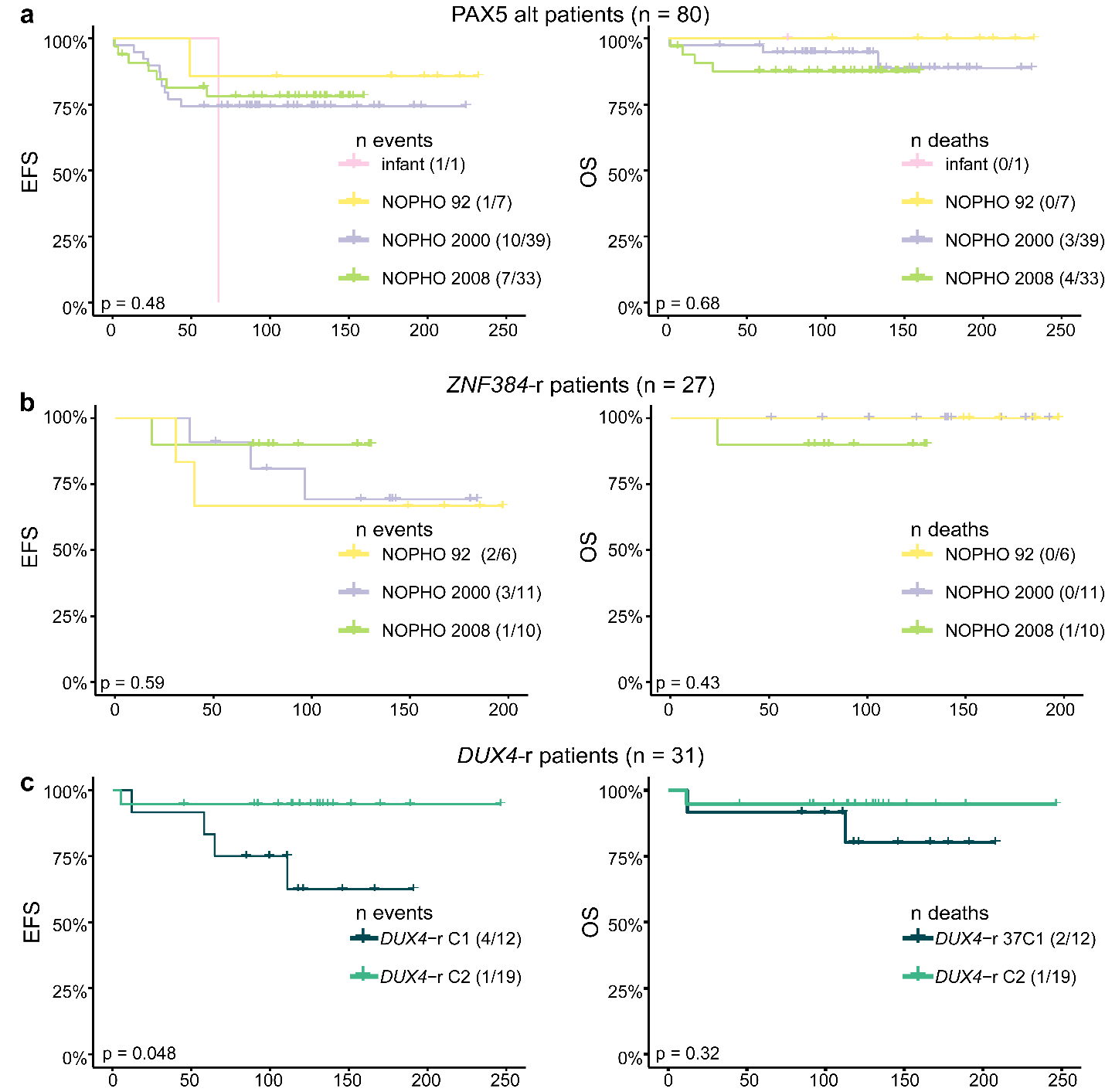

**S15. Survival analysis after B-other re-classification.** Kaplan-Meier survival curves depicting Event Free Survival (left) and Overall Survival (right). a) PAX5alt/dic(9;20) at diagnosis (n = 30) and re-classified PAX5alt (n = 50) stratified by treatment protocol before and after MRD introduction in the NOPHO-2008 protocol. b) ZNF394-r (n = 27) stratified by treatment protocol before and after MRD introduction in the NOPHO-2008 protocol. c) DUX4-r GEX-driven sub-clusters (n = 31) based on the GEX levels of the 425 ALLIUM GEX genes.

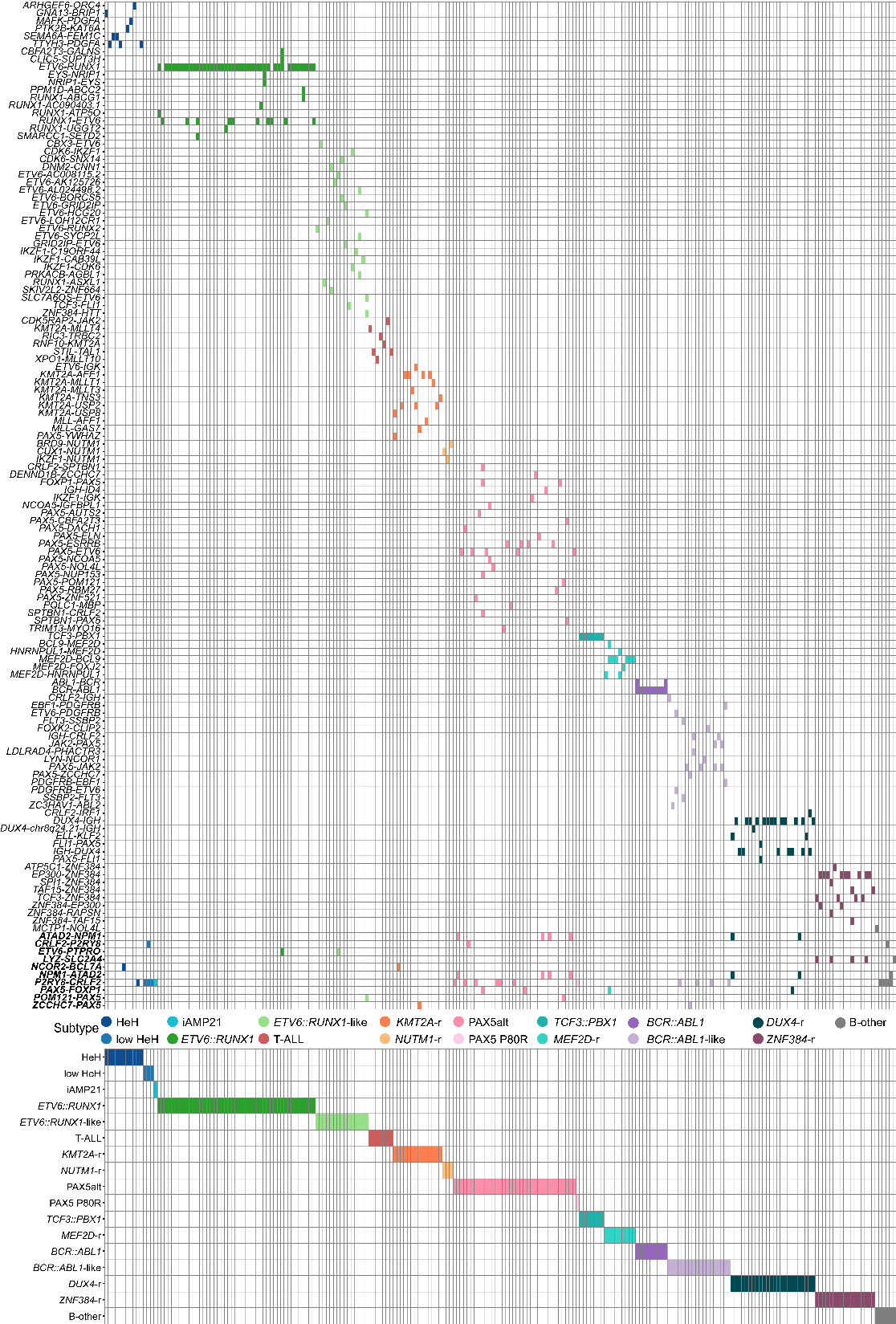

**S16. Fusion gene distribution across 225 ALL patients after molecular reclassification.** The patients are denoted in columns and fusion genes in rows**.** In total 131 unique fusion genes (including the reciprocal cases) were detected. Ten fusion genes appeared across multiple subtypes (denoted with bold Italics on the bottom).

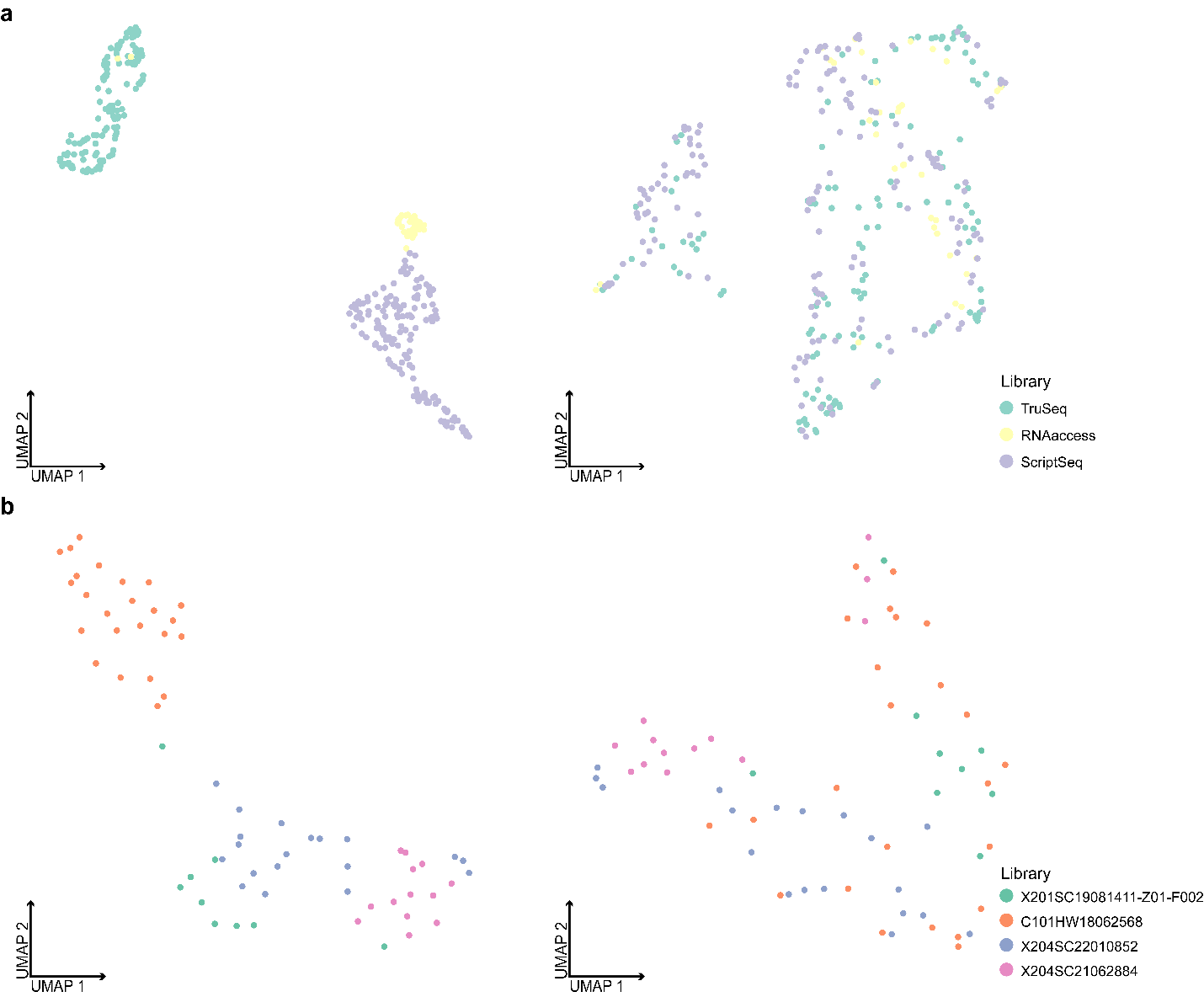

**S17. Unsupervised dimensionality reduction with UMAP before and after correcting for RNA-seq batch effects.** a) UMAP plots for UMAPs 1 and 2, demonstrating all patient samples (n = 328) in our GEX dataset before (left panel) and after (right panel) batch correction for the three preparation libraries. b) UMAP plots for UMAPs 1 and 2, demonstrating all patient samples (n = 65) in the NOPHO-Finland dataset before (left panel) and after (right panel) batch correction for the four preparation libraries.
