## Supplementary Methods and Results for "Multimodal classification of molecular subtypes in pediatric acute lymphoblastic leukemia"

*Corresponding author:

Dr. Jessica Nordlund

Box 1432, BMC

75144 Uppsala, Sweden

### Supplementary Materials and Methods

#### Control samples

Non-leukemic control samples were used to identify samples with potential low blast count. 450k array DNA methylation (DNAm) data generated from CD19+ B-cells, CD3+ T-cells from healthy blood donors, and bone marrow/peripheral blood from ALL patients in remission were downloaded from GSE49031. RNA Sequencing (RNA-seq) data from CD19+ B-cells, CD3+ T-cells from healthy blood donors, and two ALL patient samples in remission were obtained from a previous study ^1^. ALL control samples were processed as described in the **Material and Methods** for the ALL samples.

#### External Validation datasets

External data sets were used to further validate ALLIUM DNAm and GEX: 1) A beta-value matrix based on 450k array derived from 227 pediatric ALL patients were obtained from the GEO entry GSE56600^2^. 2) A GEX dataset based on RNA-seq from 19 pediatric ALL of known subtype was retrieved from GEO (GSE161501)^3^. First, the gene count matrix downloaded from GSE161501 was lifted over from the hg19 reference genome to GRCh38.103 gene annotation file based on ENSEMBL IDs. All missing genes were replaced with zeros. The data were filtered and normalized using the workflow described in the Materials and Methods. 3) RNA-seq data from 65 BCP-ALL patients were obtained from GEX NOPHO**–**Finland. The raw sequencing data were processed using the workflow described in the **Materials and Methods**.

#### Nearest Shrunken Centroid Classifiers

The Nearest Shrunken Centroid (NSC) ^4^ classification approach was selected due to its ability to perform training and feature selection (FS) based on a hyperparameter called *shrink threshold*. The shrink threshold moves each class’ centroid (mean) for every feature (gene or CpG site) towards zero. Features with zero or constant value across all classes (non-informative features) are not used during the prediction procedure. The goal of this workflow is to remove noise from the data and capture meaningful signatures for each subtype.

##### Classifier Architecture during optimization

The model was trained on the following 12 ALL groups with one or more subtype members and a control classifier comprising of healthy samples: aneuploidy (HeH, low HeH, hypodiploidy and iAMP21), *ETV6* gene rearrangements (w*ETV6::RUNX1* and *ETV6::RUNX1*-like), the Philadelphia (ph) chromosome (*BCR::ABL1* and *BCR::ABL1*-like), *TCF3::PBX1*, PAX5alt, PAX5 P80R, *KMT2A*-r, *ZNF384*-r, *DUX4*-r, *MEF2D*-r, *NUTM1*-r and T-ALL.

###### Subtype groups and one-vs-rest approach

For each subtype/group classifier, a one-vs-rest approach was implemented, where all other subtypes than the one of interest formed the ‘rest’ group (**Supplementary Figure S2**). This architecture allowed to select group specific signatures which separate each subtype group from all the rest. At the end of each group loop, a set of group-specific signatures (CpG sites/genes) were obtained. The rationale behind dividing a multiclass problem into multiple binary (one-vs-rest) classification tasks on the subtype group level was to identify patients who do not belong to any class or belong to multiple ones. This is in contrast to a multiclass model, which forces all patients into a single subtype, even if they do not adequately match the subtypes used to design the classifier.

###### Group Members

If a group contained two members (two subtypes), a one vs one approach was applied to obtain member signatures that separate one from the other, i.e for differentiating between *ETV6::RUNX1* vs *ETV6::RUNX1*-like within the *ETV6*-group and *BCR::ABL1* vs *BCR::ABL1*-like for the ph-group. Otherwise, for groups with >2 members (i.e. The aneuploidy group) a one vs rest approach was implemented instead, leading to signatures that separate the member subtype from all the rest: HeH vs rest aneuploidy, low HeH vs rest aneuploidy, hypodiploidy vs rest aneuploidy, and iAMP21 vs rest aneuploidy (**Supplementary Figure S2**).

###### Classifier Optimization and Signature Selection

The training loop consisted of two cross validation (CV) loops. The outer loop performed stratified 5-fold CV repeated 5 times, resulting in 25 different train and test sets. Stratified 5-fold CV repeated 5 times took place in the inner loop as well, where each of the 25 training datasets were divided into 25 inner train and validation sets (**Supplementary Figure S2**). Stratified folds were preferred since they retained each dataset’s characteristics in terms of patient representation in each subtype group after splitting the datasets into k-folds.

The DNAm dataset underwent pre-processing during model optimization. More specifically, CpG sites with >10% missing values across all patient samples were discarded, as well as sites with average b-value class-wise pair difference < 20%. Furthermore, the data were imputed for missing values using the median. These pre-processing steps took place iteratively on the outer loops by using the 25 outer train sets (**Supplementary Figure S2**). Then, the same filtering and imputation was applied on the test tests during CV.

This iterative process was repeated for each of the shrink threshold parameter values. The shrink threshold parameters varied between subtypes as the number of patients within each group varied as well (**Supplementary Table S26-S27**), and applied only during model optimization to obtain the best set of features per threshold value. In the inner loop, the model was trained using the inner train sets with the selected shrink threshold and all informative features per fold were obtained. At the end of all 25 inner folds, all features appeared on all 25 inner folds were kept, as a feature across all folds had a higher chance of holding significant information. The model was then trained on the outer train sets with the corresponding top features per outer fold, tested on the test sets and the test F1 score per outer fold was obtained. Finally, the consensus features were selected based on the highest test F1 score across all 25 outer folds (**Supplementary Figure S2**).

Additionally, external CV took place following the same architecture as previously described for optimization using the consensus signatures selected for each shrink threshold to obtain train, test and CV F1 scores. The data was first fitted on the inner train sets to obtain the mean CV F1 scores across all inner folds per outer fold, and then on the outer train sets to obtain both train and test F1 scores per outer fold. Finally the mean train, CV and test F1 scores across all 25 outer folds per shrink threshold were kept (**Supplementary Figure S2**). Then, in a final feature selection step the signatures of the threshold with the best performing model, in terms of classification overall F1 scores, were selected for each classifier. Since the difference in F1 scores was very small across shrink thresholds, a manual rule was set, where if applicable, to choose the shrink threshold that included the highest number of features but not more than 40 genes or CpGs per subtype group. At the end, all signatures appearing in more than one subtypes were dropped to retain only the unique subtype-specific ones.

##### Classifier Architecture during training

###### Groups and members

Each classifier was trained either using its own signatures (DNAm) or all signatures (GEX) selected by the final feature selection step, as the DNAm and GEX classifiers performed better with subtype specific or all signatures approaches respectively. The same group-member strategy as the one mentioned during optimization was used. A one-vs- rest approach was implemented for the groups as previously described and a one-vs-one to separate one member from the other (e.g. t(12;21)*ETV6-RUNX1* vs *ETV6-RUNX1*-like), if the group contained two subtype members. In contrast, an overall multiclass classifier was selected for the aneuploidy members, as the subtypes were mutually exclusive there was no need for a one vs rest approach to be implemented (HeH, low HeH, iAMP21 and hypodiploid (only on the DNAm)) (**Supplementary Figure S3**).

##### Classification Predictions

The first part of the prediction loop consisted of the group prediction. Each sample was be tested on all 12 group classifiers and healthy classifier, and when the results highlighted aneuploidy, ETV6 or ph-group, the process continued further. Since a high group score was obtained, it was upon to the model to decide whether a sample belonged to one of the member subtypes (**Supplementary Figure S3**). The generated results provided information both in group and subtype level. The final subtype decisions were made based on the group probability scores. Each sample could receive one of the following labels: no class if ‘other’ was predicted on all group classifiers, multi-class if > 1 classes were assigned, or single class. To handle the multi-class cases, the group with the highest probability score was picked as the final group prediction, and the subtype member with the highest score was obtained to give the final subtype prediction (**Supplementary Figure S3**). A manual check flag was raised if the probability score of the final prediction was <70%.

#### Performance Metrics

The classification performance metrics used included sensitivity and recall (True Positive Rate, TPR (1)), specificity (True Negative Rate, TNR (2)) and precision (Positive Predictive Value, PPV (3)). Balanced accuracy (4) was used instead of standard accuracy, since the data was imbalanced. High accuracy scores might be misleading as the performance of the under-represented class could be poor. Balanced accuracy mitigates this effect by giving an overall score taking imbalance into consideration. F1- score (5) represents the harmonic mean between specificity and precision. The closer to 1 all scores are, the better the model performs.

$$Sensitivity=\frac{TP}{TP+FN} (1)$$

$$Specificity=\frac{TN}{TN+FP} (2)$$

$$Precision=\frac{TP}{TP+FP} (3)$$

$$Balanced accuracy=\frac{Sensitivity+Specificity}{2} (4)$$

$$F1 score=2 \times\frac{Sensitivity \times Precision}{Sensitivity+Precision} (5)$$

Finally, during model optimization to obtain the optimal signature set based on performance, F1 weighted score (6) was utilized since both comparing groups were equally important. Our classifiers did not use control (negative) vs disease state (positive) samples, thus it was of high importance to maintain good performance on both comparing groups, either it was one subtype vs all the rest, or one subtype vs another. F1 weighted score takes into consideration all groups and calculates each F1 score multiplied by a weight (w) based on group sample size to avoid issues due to class imbalance and then averages it.

$$F1 weighted=\frac{1}{N}\sum_{i=1}^{N} {F1}_{i} \times w_{i}(6)$$

### Supplementary Results

#### Internal validation for ALLIUM GEX

We internally created a validation set to evaluate ALLIUM GEX, which consisted of 13 samples from 11 unique patints (12 samples with known subtype, 1 B-other). The samples were either taken at ALL relapse (n = 8) or a new library was prepared and re-sequenced (n = 5). Ten of the 12 (83.3%) predictions matched with the true ALL subtype (**Supplementary Figure S4**). Eleven of the 13 predictions, in total, were concordant (**Supplementary Table S9, S10**). The samples with mismatches include ALL_1026 (PAX5alt) and ALL_257 (*ZNF384*-r), which were taken at relapse and predicted as “no class” by ALLIUM GEX. Of interest, patient ALL_257 (*ZNF384*-r) was predicted as *ETV6::RUNX1*-like for the diagnostic sample. ALL_257 is known to carry characteristics of both subtypes *ZNF384*-r and *ETV6::RUNX1*-like ^1,5^.

#### External validation for ALLIUM DNAm: GSE56600

GSE56600 contains 450k array DNA methylation data for 227 BCP-ALL patients. ALLIUM DNAm correctly predicted 84.2% (112/133) of the patients with known subtype from this cohort. From the remaining 21 misclassified samples, 13 were classified as “control”, suggesting low blast counts, and in the last sample “no class” was predicted (**Supplementary Figure S4, Supplementary Table S11**). This cohort contained 94 patients with B-other subtype, of these ALLIUM DNAm classified 68 (72.3%) to a unique subtype, 16 with no class prediction (17.0%), and 10 as “control”/low blast count (10.6%) (**Supplementary Table S11**). The classifier showed an overall sensitivity and specificity of 79.3% and 99.1% respectively (**Table 2, Supplementary Table S14**).

#### External validation for ALLIUM GEX: GSE161501

GSE161501 contained RNA-seq data for 19 BCP-ALL patients. ALLIUM GEX classifier correctly classified the subtype of each of the 19 patients in this dataset (**Supplementary Figure S4, Supplementary Table S12**), with an overall sensitivity and specificity of 100% and 100% respectively (**Table 2, Supplementary Table S14**).

#### External validation for ALLIUM GEX: NOPHO-Finland

RNA-seq data were available for 65 BCP-ALL samples. In total, 55 were of known subtype and 96.4% (n = 53) were correctly classified (**Supplementary Figure S4, Supplementary Table S13**). The nine B-other samples were all assigned to a subtype: *DUX4*-r (n = 3), *ETV6::RUNX1*-like (n =2), *ZNF384*-r (n = 2), *MEF2D*-r (n = 1) and iAMP21 (n =1). For patient ALLT-351, with hypodiploid subtype, ALLIUM GEX returned “no class” as ALLIUM GEX was not trained on this subtype. The model performance for the samples of known subtype demonstrated an overall sensitivity and specificity of 99.3% and 99.8% respectively (**Table 2, Supplementary Table S14**).
